# Expression of γ-globin genes in β-thalassemia patients treated with sirolimus: results from a pilot clinical trial (Sirthalaclin)

**DOI:** 10.1101/2021.12.20.21267830

**Authors:** Cristina Zuccato, Lucia Carmela Cosenza, Matteo Zurlo, Jessica Gasparello, Chiara Papi, Elisabetta D’Aversa, Giulia Breveglieri, Ilaria Lampronti, Alessia Finotti, Monica Borgatti, Chiara Scapoli, Alice Stievano, Monica Fortini, Eric Ramazzotti, Nicola Marchetti, Marco Prosdocimi, Maria Rita Gamberini, Roberto Gambari

## Abstract

**Introduction:** The β-thalassemias are due to autosomal mutations of the β-globin gene, inducing absence or low-level synthesis of β-globin in erythroid cells. It is widely accepted that high production of fetal hemoglobin (HbF) is beneficial for β-thalassemia patients. Sirolimus, also known as rapamycin, is a lipophilic macrolide isolated from a strain of Streptomyces hygroscopicus found to be a strong HbF inducer in vitro and in vivo. In this study, we report biochemical, molecular and clinical results of the sirolimus-based NCT03877809 clinical trial (A Personalized Medicine Approach for β-thalassemia Transfusion Dependent Patients: Testing sirolimus in a First Pilot Clinical Trial: Sirthalaclin).

**Methods:** Accumulation of γ-globin mRNA was analyzed by reverse-transcription-quantitative PCR and the hemoglobin pattern by HPLC. The immunophenotype was analyzed by FACS using antibodies against CD3, CD4, CD8, CD14, CD19, CD25.

**Results:** The results were obtained in 8 patients with β+/β+ and β+/β0 genotypes, treated with a starting dosage of 1 mg/day sirolimus for 24-48 weeks. The first finding of the study was that expression of γ-globin mRNA was increased in blood and erythroid precursor cells isolated from β-thalassemia patients treated with low-dose sirolimus. A second important conclusion of our trial was that sirolimus influences erythropoiesis and reduces biochemical markers associated to ineffective erythropoiesis (I.E.) (excess of free α-globin chains, bilirubin, soluble transferrin receptor and ferritin). In most (7/8) of the patients a decrease of the transfusion demand index was observed. The drug was well tolerated with minor effects on immunophenotype, the only side effect being frequently occurring stomatitis.

**Conclusions:** The data obtained indicate that sirolimus given at low doses modifies hematopoiesis and induces increased expression of γ-globin genes in a sub-set of β-thalassemia patients. Further clinical trials are warranted, considering the possibility to test the drug in patients with less severe forms of the disease and exploring combination therapies.

## Introduction

The β-thalassemias are among the most common inherited hemoglobinopathies worldwide, and are due to autosomal mutations in the gene encoding β-globin, causing an absence or low-level synthesis of this protein in erythropoietic cells [1-3]. The phenotypes range from asymptomatic (β-thalassemia trait or carrier) to clinically relevant anemia which is categorized as transfusion-dependent β-thalassemia (TDT, including thalassemia major) and non-transfusion-dependent β-thalassemia (NTDT, thalassemia intermedia).

About 80 to 90 million people (∼ 1.5 % of the global population) are carriers of β-thalassemia with approximately 60,000 symptomatic individuals born annually [4-7]. Worldwide, nearly 300 mutations around the β-globin gene have now been reported to cause β-thalassemia [8]. Most are point mutations either in the gene or its immediate flanking regions, but some are caused by small deletions removing all or part of the β-globin gene.

It is widely accepted that high production of fetal hemoglobin (HbF) is beneficial for β-thalassemia patients [9-11]. The earliest clinical observations suggesting the advantageous role of HbF in thalassemia came from patients with rare forms of β-thalassemia, particularly those with large deletions responsible of δβ^0^-thalassemia or hereditary persistence of fetal hemoglobin (HPFH), with absence of β-globin production but associated to high quantity of γ-globin chain production, resulting in high levels of HbF with a relatively benign clinical course [12]. More recent clinical studies have disclosed that the naturally higher production of HbF improves the clinical course in a variety of patients with β-thalassemia [13-17]. Accordingly, these observations have promoted research for inducers of HbF that can therapeutically reproduce what occurs in β-thalassemia patients with natural persistence of higher levels of HbF [18-21]. Furthermore, approaches using genome editing are available finalized to induction of HbF following elimination of genomic sequences encoding for transcriptional repressors or genomic sequences targeted by these regulatory factors [22-25]. In this context clinical trials are occurring, such as NCT03655678 (A Safety and Efficacy Study Evaluating CTX001 in Subjects With Transfusion-Dependent β-Thalassemia), based on the use of autologous CRISPR-Cas9 Modified CD34+ Human Hematopoietic Stem and Progenitor Cells (hHSPCs) using CTX001 [26].

Sirolimus, also known as rapamycin [27] is a lipophilic macrolide isolated from a strain of *Streptomyces hygroscopicus* found in a soil from Easter Island (known by the inhabitants as Rapa Nui). Sirolimus has been shown to be a strong inducer of HbF in vitro [28-31], in vivo using animal model systems [32-34] and in some patients affected by sickle-cell disease. Nowadays this drug is used as immunosuppressant in combination with other drugs (e.g. cyclosporine or corticosteroids) in the setting of prevention of transplant rejection [35,36] (see **Supplementary Figure S1** for an historical summary of key actions and findings concerning sirolimus).

The relevance of these reports is due to the fact that no treatment has been so far approved concerning HbF inducers in thalassemia. While hydroxyurea (HU) is frequently used (despite the lack of formal approval), its use is limited by the potential adverse effects and reported efficacy in only a subset of patients [37-39]. Thus, a substantial percentage of thalassemia patients are not treated with HU and may benefit by other treatments. Moreover, some patients become HU resistant after long term treatment [40].

The working hypothesis to propose sirolimus for β-thalassemia is that the reported sirolimus-dependent increase of the HbF levels will improve the clinical status of the β-thalassemia patients, with the aim of reducing the transfusion need in transfusion dependent patients, the majority of the population present in Europe. As far as transfusion independent patients, sirolimus effects should be studied with end points partly different from the ones we will describe in this manuscript.

On the basis of the published information about the effects of sirolimus on HbF, the Orphan Drug Designation (ODD) has been obtained by Rare Partners for this repurposed drug from the European Medicines Agency (EMA) for the treatment of β-thalassemia (code EU/3/15/1585) and of SCD (code EU/3/17/1970). ODD of sirolimus as HbF inducer in β-thalassemia and SCD was also obtained by the U.S. Food and Drug Administration (FDA) (see **Supplementary Figure S1**).

In this manuscript, we report the major biochemical, molecular and clinical outcomes of the NCT03877809 clinical trial (A Personalized Medicine Approach for β-thalassemia Transfusion Dependent Patients: Testing sirolimus in a First Pilot Clinical Trial: Sirthalaclin), focusing on patients with β^+^-thalassemia genotypes [41]. This trial was based on the use of low-dosages of the repurposed drug sirolimus (rapamycin) for a 12 months period [42,43].

The main objective of this interventional, pilot, open-label phase II study with sirolimus in patients with β-TDT (transfusion-dependent thalassemia) was to verify its efficacy as in vivo HbF inducer aiming to reduce transfusions need with an overall good tolerability. A second objective was to verify possible occurrence of adverse events (AEs) during the clinical trial, resulting from any abnormal laboratory or instrumental finding, symptom or disease temporally associated with the use of the investigational product.

## 2. Methods

### Patient recruitment and sirolimus treatment

Recruitment of the Sirthalaclin trial (EudraCT n° 2018-001942-33, NCT 03877809) was at the Thalassemia Centre of Azienda Ospedaliera-Universitaria S.Anna (Ferrara, Italy). The β-thalassemia patients have been recruited among patients with β^+^/β^+^ and β^+^/β^0^ genotypes. The study was approved by Ethical Committee in charge of human studies at Arcispedale S. Anna, Ferrara (release of the approval: November 14, 2018). The patients who fulfill inclusion/exclusion selection criteria (enlisted in the **Supplementary Tables S1** and **S2**) received a patient information sheet to read and time to clarify doubts with investigators before consenting. Informed written consent from all participants was obtained before recruiting them into the study.

The investigational drug in the form of coated tablets (0.5 mg sirolimus) has been provided to the patients in blisters, adequately labelled with the information regarding the study. At visit 2 (V2, day 0, see **Figure 1**) patients were instructed to take 2 tablets (0.5 mg each). The sirolimus blood level was measured for the first time after 10 days of therapy and then on day 90 (visit 6, V6), on day 180 (visit 8, V8), on day 240 (visit 9, V9) and on day 360 (visit 11, V11). The starting sirolimus dosage was 1 mg/day. Treatment dose was planned to be increased up to 2 mg/day or decreased to 0.5 mg/day in order to maintain sirolimus whole blood concentration in a range between 5-8 ng/ml. The dosage has been kept constant unless a reduction dictated by side effects. All other standard treatments, including blood transfusions and iron chelation therapy have been continued. All patients received leuco-depleted packed red blood cell transfusions in order to maintain a pre-transfusion level of hemoglobin between 9 and 10.5 g/dl, in agreement with the International Thalassaemia Federation (T.I.F.) Guidelines [44].

**Figure 1.**
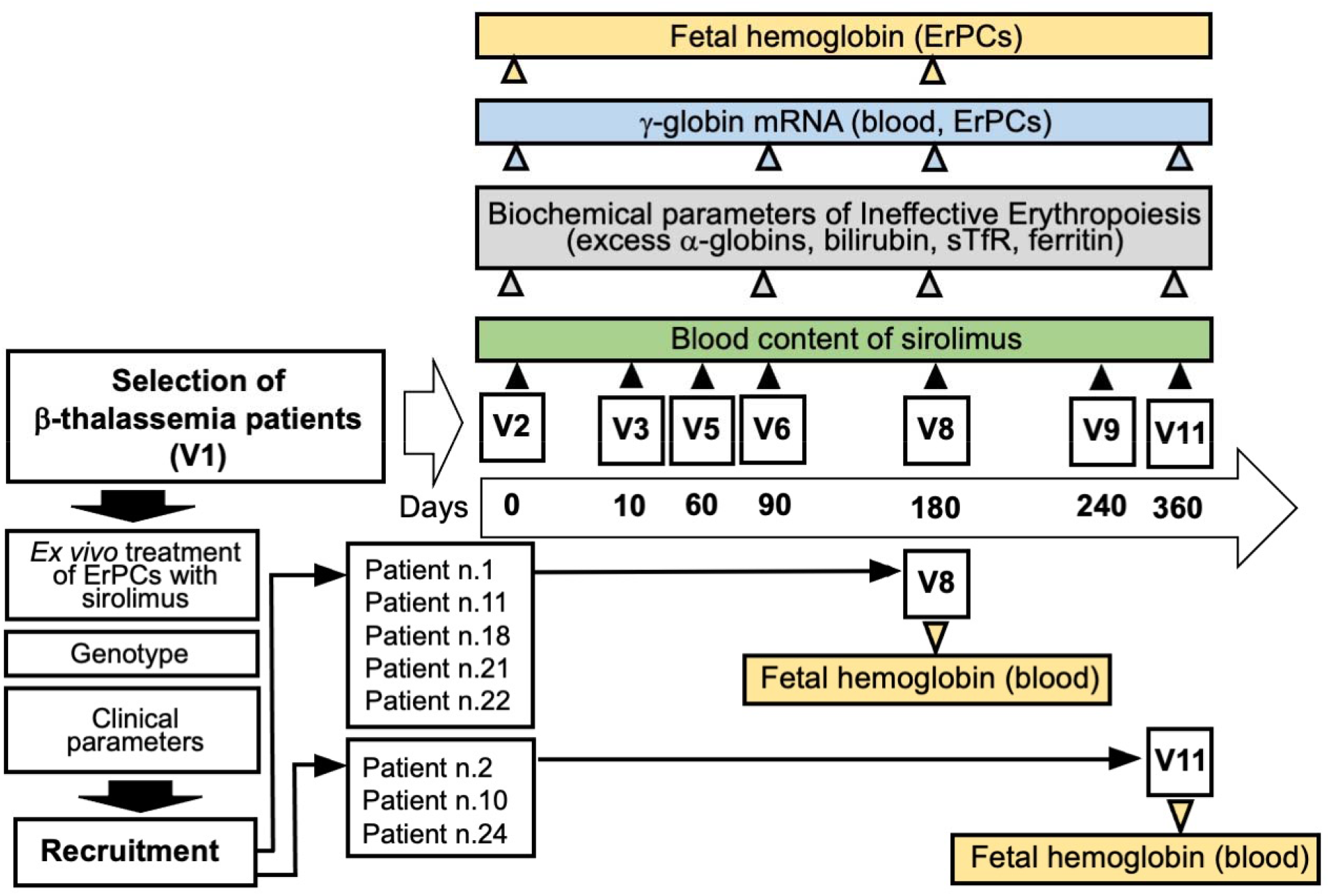
Flow-chart summarizing the NCT03877809 Sirthalaclin clinical trial and the key analyzed presented in the present study. Five patients concluded the trial at V8 (180 days) as indicated. Fetal hemoglobin and excess of free α-globin chains were analyzed by HPLC. Content of γ-globin mRNA was analyzed by RT-qPCR. The scheme of the trial was elsewhere described [42].

### Analysis of the sirolimus blood content

The analysis of the sirolimus blood content in thalassemic patients was carried out by *Solid-Phase Extraction* SPE *LC/MS/MS*. Briefly, 100 µl of sample/Cal/QC blood samples from thalassemic patients were added to 200 µL of deproteinizing solution (ZnSO4: Methanol); the obtained solutions were vortexed for 10 sec and centrifuged at 13000 rpm for 5 minutes; the supernatants were recovered and transferred to the dedicated microplates to be housed in the mass tandem autosampler (Series 200 AS). The LC/MS system consists of an HPLC Perkin-Elmer spa coupled with a Mass Spectrometer Ab-Sciex API 2000 (AB Sciex LLC, Redwood City, CA, USA). The injected sample (15 μl) goes through a cleaning phase via online SPE thanks to the use of the preparative POROS R1/20 column. The samples were then separated by analysis on Luna Phenyl-Hexyl 50 × 2.1 mm, 5 μm (Phenomenex) analytical column. The analyses were conducted at a temperature of 60°C, with a duration of about 3.2 minutes at a flow of 0.4 mL/min and according to the gradient below: from 0 to 1 min the poros is charged to 0% of mobile phase B; from 1 to 1.01 a climb up to 100% of B; 100% of B is maintained up to 2.8 min. The analysis ends at 3.2 by rebalancing the system at 0% of B. For the mass acquisition, the instrument uses the ionization mode via ESI (electrospray ionization) in positive polarity, the electrode voltage is set at 5500 V and the capillary temperature set at 275°C. The mass acquisition mode is MRM (Multiple Reaction monitoring) identifying as characteristic transitions for the Siro and Ever-d4 respectively 931.6/864.6 and 981.6/914.4.

### In vitro culture of Erythroid Progenitors (ErPCs) from β-thalassemia patients

The two-phase liquid culture procedure was employed as previously described [29, 45-47]. Mononuclear cells were isolated from peripheral blood samples of β-thalassemia patients by Ficoll-Hypaque density gradient centrifugation. After isolation, the mononuclear cell layer was washed three times by adding 1x phosphate-buffered saline (PBS) solution and seeded in α-minimal essential medium (α-MEM, Sigma Genosys) supplemented with 10% FBS, (Celbio, Milano, Italy), 1 µg/ml cyclosporine A (Sandoz, Basel, Switzerland), 10% conditioned medium from the 5637 bladder carcinoma cell line culture and stem cell factor (SCF, Life Technologies, Monza, MB, Italy) at the final concentration of 10 ng/ml. The cultures were incubated at 37°C, under an atmosphere of 5% CO2. After 7 days in this phase I culture (at 37°C, under an atmosphere of 5% CO2 in air, with extra humidity), the non-adherent cells were harvested from the flask, washed in 1x PBS, and then cultured in phase II medium, composed of α-MEM medium, 30% FBS (Celbio), 1% deionized bovine serum albumin (BSA) (Sigma Genosys), 10^−5^ M β-mercaptoethanol (Sigma Genosys), 2 mM L-glutamine (Sigma Genosys), 10^−6^ M dexamethasone (Sigma Genosys) and 1 U/ml human recombinant erythropoietin (EPO) (Tebu-bio, Magenta, Milano, Italy) and stem cell factor (SCF, BioSource International, Camarillo, CA, USA) at the final concentration of 10 ng/ml. Erythroid precursor cells differentiation was assessed by benzidine staining [29, 45-46]. Sirolimus was administered to ErPCs at 100 nM concentration.

### RNA extraction from blood and ErPCs

The total cellular RNA was extracted from blood and ErPCs by using TRIzol™ LS (Invitrogen) and TRI Reagent® (Sigma-Aldrich, Saint Louis, Missouri, USA), respectively and following the manufacturer’s instructions. The protocol used for extraction of whole blood RNA was strictly followed with these steps: (a) 100 μl of whole blood were diluted in 3 volumes of 1xPBS and processed using 1 ml of TRIZOL LS (added without reverse pipetting in order to avoid any clumping of blood); (b) the isolated RNA was washed once with cold 75% ethanol, dried and dissolved in 10-20 μl nuclease free water before use. The procedure for RNA extraction from ErPCs was similar to that described for blood. In this case 800 μl of TRI Reagent® were added to a dry pellet of 4-6 × 10^6^ cells.

### RT-qPCR analysis of γ-globin gene expression

For gene expression analysis 500 ng of total RNA was reverse transcribed by using the TaqMan® Reverse Transcription Reagents and random hexamers (Applied Biosystems, Life Technologies, Carlsbad, CA, USA). Quantitative real time PCR assay, to quantify the expression of the globin genes, was carried out using two different reaction mixtures, the first one containing γ-globin probe and primers, the second one containing GAPDH, RPL13A, β-actin probes and primers. The primers and probes used are listed in **Table I**.

**TABLE I.**
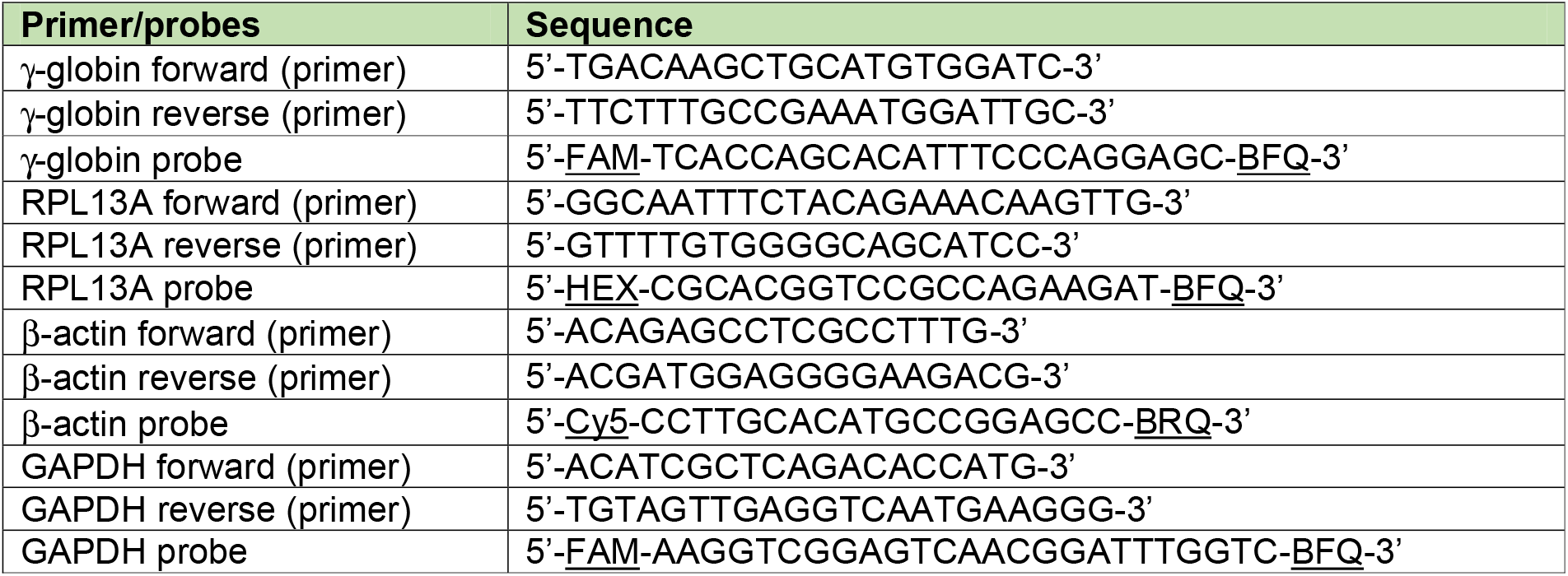
Sequences of the primers and probes employed.

Each reaction mixture contained 1x TaKaRa Ex Taq® DNA Polymerase (Takara Bio Inc., Shiga, Japan), 500 nM forward and reverse primers and the 250 nM probes (Integrated DNA Technologies, Castenaso, Italy). The assays were carried out using CFX96 Touch Real-Time PCR System (Bio-Rad, Hercules, California, USA). After an initial denaturation at 95°C for 1 min, the reactions were performed for 50 cycles (95°C for 15 sec, 60°C for 60 sec). Data were analyzed by employing the CFX manager software (Bio-Rad, Hercules, California, USA). To compare gene expression of each template amplified, the ΔΔCt method was used [29,30].

### HPLC analysis of hemoglobins

To evaluate the effectiveness of treatment HPLC analysis was carried using both blood and ErPCs. The ErPCs were centrifuged at 8000 rpm for 8 minutes and washed with PBS (Phosphate buffered saline). The pellets were then resuspended in a predefined volume of water for HPLC (Sigma-Aldrich, St. Louis, Missouri, USA). This was followed by three freeze/thaw cycles on dry ice in order to lyse the cells and obtain the protein extracts. Lysates were centrifuged for 5 min at 14,000 rpm and the supernatant was collected. Hemoglobin analysis was performed by loading the protein extracts into a PolyCAT-A cation exchange column and then eluted in a sodium-chloride-BisTris-KCN aqueous mobile phase using HPLC Beckman Coulter instrument System Gold 126 Solvent Module-166 Detector which allows us to obtain a quantification of the hemoglobins present in the sample. The reading is performed at a wavelength of 415nm, and a commercial solution of purified human HbF (Analytical Control Systems Inc, IN, USA) extracts has been used as standard. The values thus obtained are processed using “32 Karat software”.

### Calculation methods for the transfusion indexes

Transfusion episodes occurred in two periods, prior to sirolimus treatment (about 180 days before V2) and under sirolimus treatment (from V2 to V8) were considered. For each period, transfusion indexes were calculated, including the average pre-transfusion hemoglobin concentration (g/dL) and the red cell consumption (mL of pure red cells per kg body weight per year). The index of transfusion demand was calculated dividing the red cell consumption by the average pre-transfusion hemoglobin concentration. If the endogenous production of hemoglobin was increases under sirolimus treatment, then this parameter was expected to decrease proportionally. Detail and a representative example for the calculations of transfusion indexes have been included in **Supplementary materials**.

The analysis of erythroblast count, reticulocytes count and total bilirubin, lactate dehydrogenase (LDH), soluble transferrin receptor (sTfR), erythropoietin and ferritin levels have been performed using standard clinical assays at the Laboratory of Analysis, Arcispedale S. Anna di Ferrara, Ferrara, Italy.

### Analysis of the immunophenotype

Concerning the immunophenotype analysis performed by flow cytometry, peripheral blood mononuclear cells (PBMCs) were isolated from whole blood of recruited patients during different time-points (V2-V6-V8-V11). Briefly, a small amount of blood was layered on ficoll and centrifuged to collect the buffy coat (mainly white blood cells, ErPCs and platelets), then a series of washes in PBS are needed to clean the cells and eliminate the platelet component. At the end of the process the isolated PBMCs were frozen in several cryo-vials using FBS with addiction of 10% DMSO as a cell cryopreservation medium and stored in liquid nitrogen. At the end of the Clinical Trial, for each patient, we thawed one cryo-vial of each time-point in order to perform the immunophenotype analysis. Cells were thawed and centrifuged in RPMI medium and then washed in PBS and counted: one million cells were selected for each time-point to proceed with the antibody staining. The cells were stained with LIVE/Dead™ Fixable Aqua – Dead Cell Stain Kit (from Thermo Fisher, Waltham, MA, USA) and incubated 10’ in the dark to color cells that have lost membrane integrity. After a PBS wash, cells were stained with a mixture of antibodies that target different membrane receptors (antibodies against CD3, CD4, CD8, CD14, CD19 and CD25 markers) and incubated 15’ in the dark. At the end of the staining process, cells were washed in PBS to reduce the background, resuspended in 200 μl of PBS and analyzed by flow cytometry using the BD FACSCanto II cell analyzer (Becton Dickinson) [47-49].

### Statistical analysis

All the data were normally distributed and presented as mean ± S.D. Statistical differences between groups were compared using paired t-test or one-way repeated measures ANOVA (ANalyses Of VAriance between groups) followed by LSD post-hoc tests. Statistical differences were considered significant when p< 0.05 (*), highly significant when p< 0.01 (**).

## Results

### Selection of the β-thalassemia patients and determination of the in vitro response of ErPCs to sirolimus

To all the patients who fulfilled inclusion/exclusion selection criteria (enlisted in **Supplementary Tables S1 and S2**), a patient information sheet has been given. The necessary time to discuss and clarify all doubts with investigators was allowed, before consenting. Informed written consent from all participants was obtained before recruiting the patients into the study. At the time of enrolment, information on sociodemographic background, family history, medical history, present medical problems and concomitant medication have been reviewed. In the selection period of the study physical examination, cardiac evaluation, abdominal ultrasound and blood tests were performed. Patients eligible for the study were patients who fulfilled inclusion/exclusion criteria and who were responsive to sirolimus in vitro (according to laboratory specific definition: ≥ 20% increase of HbF in comparison with samples not treated with sirolimus) were recruited.

**Figure 1** shows a pictorial flow-chart summarizing the key steps of the NCT03877809 Sirthalaclin clinical trial, including the preliminary phases allowing recruitment of β-thalassemia patients and the scheme of the biochemical and molecular assays conducted and presented in this manuscript.

As indicated in the elsewhere presented protocol [42] and in **Figure 1**, initially erythroid precursor cells (ErPCs) were treated in vitro and only patients whose cells were responsive were candidates to receive orally the drug. A total of 24 β^+^-thalassemia patients were recruited for this analysis. Representative HPLC patterns obtained after exposure to sirolimus of ErPCs isolated from patients n.9, n.14, n.15 and n.17 are shown in **Figure 2**. The arrows indicate the position of the hemoglobin peaks, including the major HbF peak (HbF_0_). The ErPCs cultures from two patients (n.9 and n.17) were not responders (increase of HbF < 20%), while the ErPCs cultures from the two other patients (n.14 and n.15) were considered responders (increase of HbF > 20%). The data obtained in ErPCs from the 24 patients studied in the selection period are reported in **Table III**. A total of 16 (66.6%) patients were found to have ErPCs responsive to the sirolimus treatment as stated in the Sirthalaclin protocol (i.e. 20% increase after in vitro sirolimus induction of the ErPCs).

**Figure 2.**
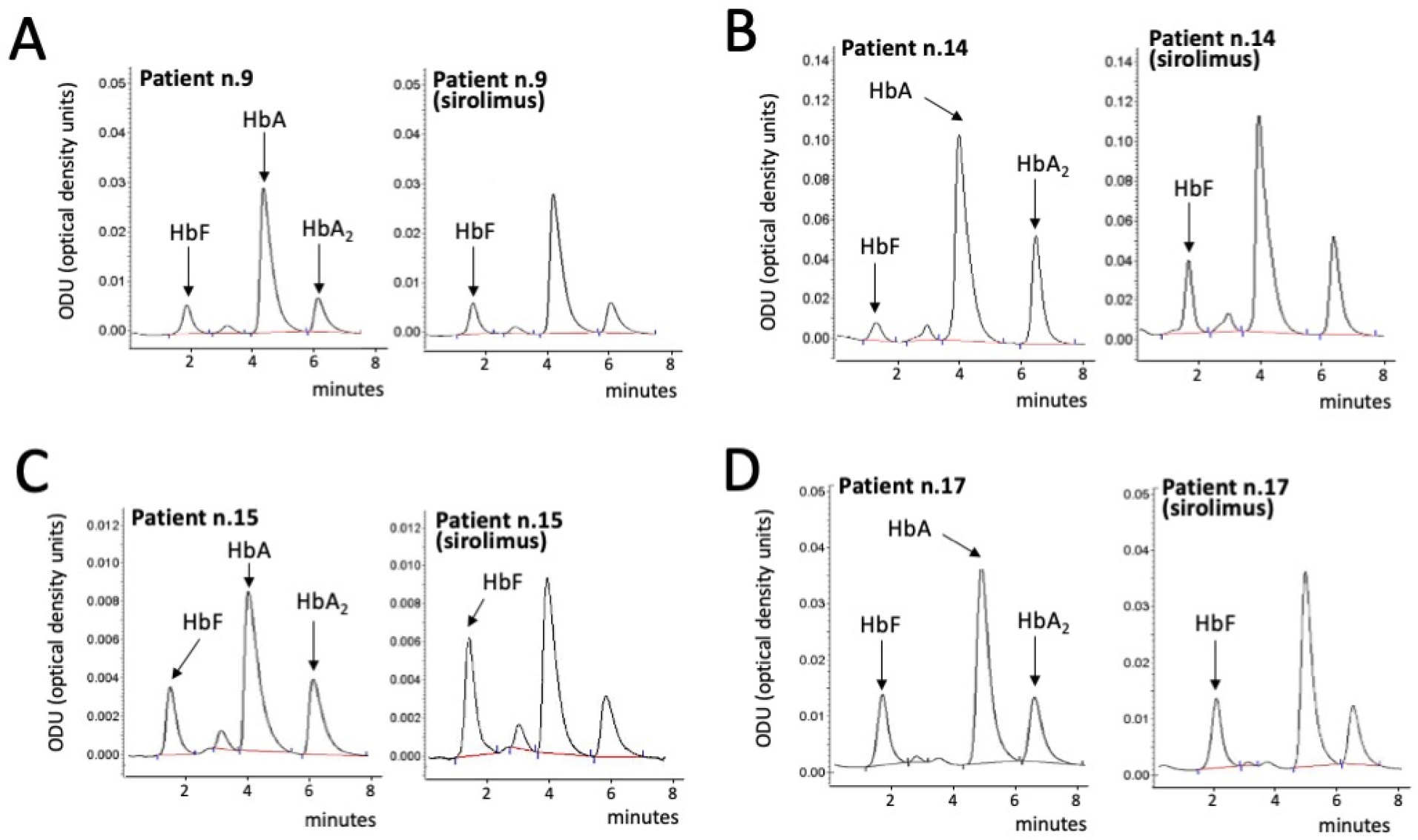
Representative HPLC patterns obtained after exposure to sirolimus of ErPCs isolated from patients n.9 (A), n.14 (B), n.15 (C) and n.17 (D). The arrows indicate the position of the HbA, HbA_2_ and HbF_0_ peaks.

**TABLE II.**
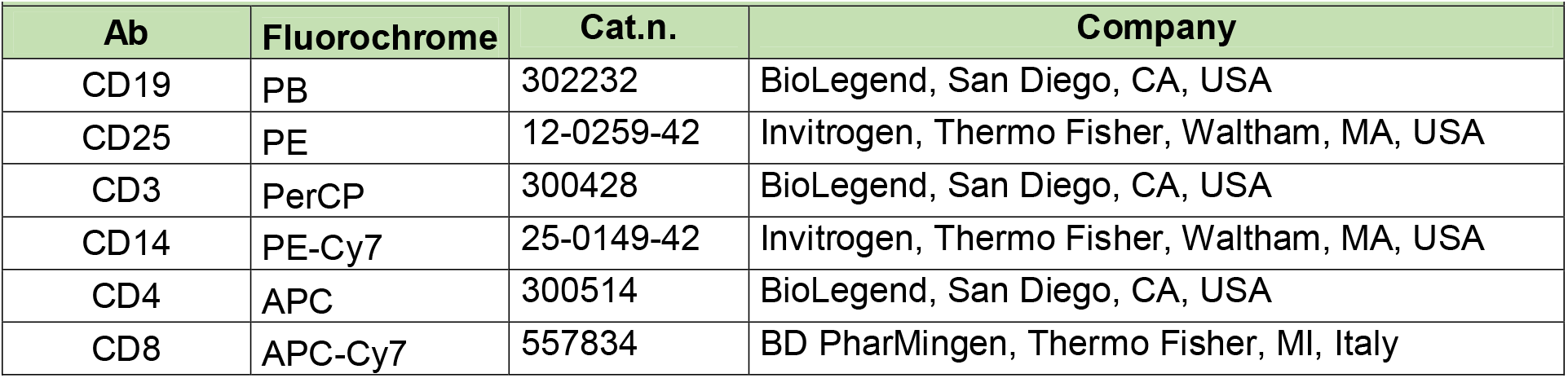
Antibodies used for immunophenotype analysis of PBMCs.

| **Ab** | **Fluorochrome** | **Cat.n.** | **Company** |
| --- | --- | --- | --- |
| CD19 | PB | 302232 | BioLegend, San Diego, CA, USA |
| CD25 | PE | 12-0259-42 | Invitrogen, Thermo Fisher, Waltham, MA, USA |
| CD3 | PerCP | 300428 | BioLegend, San Diego, CA, USA |
| CD14 | PE-Cy7 | 25-0149-42 | Invitrogen, Thermo Fisher, Waltham, MA, USA |
| CD4 | APC | 300514 | BioLegend, San Diego, CA, USA |
| CD8 | APC-Cy7 | 557834 | BD PharMingen, Thermo Fisher, MI, Italy |

**TABLE III.** In vitro response of ErPCs to sirolimus.

| **TABLE III. In vitro response of ErPCs to sirolimus** |  |  |  |  |
| --- | --- | --- | --- | --- |
| **Patient number (\*)** | **Baseline HbF (\*\*)** | **In vitro induction (% increase)** | **Recrutable** | **Treatment with sirolimus** |
| 01 | 17.80 | 21.72 | Yes | Yes |
| 02 | 10.31 | 47.40 | Yes | Yes |
| 03 | 18.15 | 29.51 | Yes | No |
| 04 | 45.09 | 11.31 | No | n.a. |
| 05 | 33.11 | 14.72 | No | n.a. |
| 06 | 17.70 | 20.28 | Yes | No |
| 07 | 9.06 | 44.92 | Yes | No |
| 08 | 17.33 | 21.20 | Yes | No |
| 09 | 12.07 | 16.30 | No | n.a. |
| 10 | 14.91 | 28.23 | Yes | Yes |
| 11 | 15.02 | 24.82 | Yes | Yes |
| 12 | 11.49 | 22.37 | Yes | No |
| 13 | 8.89 | 14.80 | No | n.a. |
| 14 | 8.32 | 61.00 | Yes | No |
| 15 | 16.74 | 61.91 | Yes | No |
| 16 | 11.10 | 16.91 | No | n.a. |
| 17 | 18.38 | 8.31 | No | n.a. |
| 18 | 21.32 | 29.11 | Yes | Yes |
| 19 | 33.18 | 10.50 | No | n.a. |
| 20 | 7.52 | 19.50 | No | n.a. |
| 21 | 9.47 | 24.71 | Yes | Yes |
| 22 | 11.24 | 29.70 | Yes | Yes |
| 23 | 19.27 | 35.11 | Yes | No |
| 24 | 14.95 | 26.31 | Yes | Yes |

(\*) The patients have been randomized in order to avoid identifying information
(\*\*) Analyzed by HPLC and expressed as % of total hemoglobins
n.a. = not applicable

Among them, 8 patients decided to start with the sirolimus intake (see **Table III)**. The age of these patients was between 40 and 60 years (4 in the range of 40-45 years; two in the range of 46-50 years; two in the range of 56-60 years). The most represented genotype was β^0^39/β^+^IVSI-110 (6 patients). One patient was β^+^IVSI-110/ β^+^IVSI-110 and one patient was β^0^39/β^+^IVSI-6. The female/male ratio was 6/2. Most of the patients (6 patients) were splenectomized. Other clinical and biochemical parameters are reported in **Supplementary Table S3**.

### Sirolimus dosage in blood

The starting sirolimus dosage was 1 mg/day with planned minimum and maximal doses of 0.5 and 2.0 mg/day respectively. The dosage was adjusted on the basis of blood levels and tolerability. The analysis of the sirolimus levels reached in peripheral blood is shown in **Table IV**. We observed high variability among patients and relatively low levels of blood sirolimus concentration in all the sirolimus-treated patients. Note that at 90 and 180 days 8 patients were taking sirolimus (average dose 1.2 and 1.7 mg/day respectively). Later on, only 3 patients remained under treatment and their blood levels were not substantially different.

**TABLE IV.** Levels of sirolimus in blood (ng/ml)

| Patient number | V2 | V3 | V5 | V6 | V8 | V9 | V11 |
| --- | --- | --- | --- | --- | --- | --- | --- |
| 1 | n.d. | 2.7 | 1.1 | 2.0 | <1.0 | (**) | (**) |
| 2 | n.d. | 2.9 | 3.7 | 2.1 | 2.0 | 5.1 | n.a. |
| 10 | n.d. | 3.1 | 2.7 | 4.5 | 2.4 | 5.9 | 4.4 |
| 11 | n.d. | 2.0 | 3.3 | 1.9 | 3.7 | (**) | (**) |
| 18 | n.d. (*) | 4.2 | 3.6 | 2.7 | 4.6 | (**) | (**) |
| 21 | n.d. | 2.1 | <1.0 | 1.7 | 3.0 | (**) | (**) |
| 22 | n.d. | 1.6 | 1.1 | 2.4 | <1.0 | (**) | (**) |
| 24 | n.d. | 2.9 | 2.1 | 1.5 | n.a. | 4.6 | n.a. (*) |

(\*) n.d. = not detected; n.a. = not available
(\*\*) Patients 1, 11, 18, 21 and 22 concluded the trial at V8

In order to compare blood levels at different time points we considered that samples with a concentration of sirolimus close to the detection limit of 1 ng/ml should be considered as containing 1 ng/ml. Using this procedure, the average values at V3, V5, V6 and V8 (**Figure 3**) were not different by ANOVA test (*p* = 0.934; **Figure 3**), indicating that repeated administration of low dose sirolimus does not cause accumulation of the drug.

**Figure 3.**
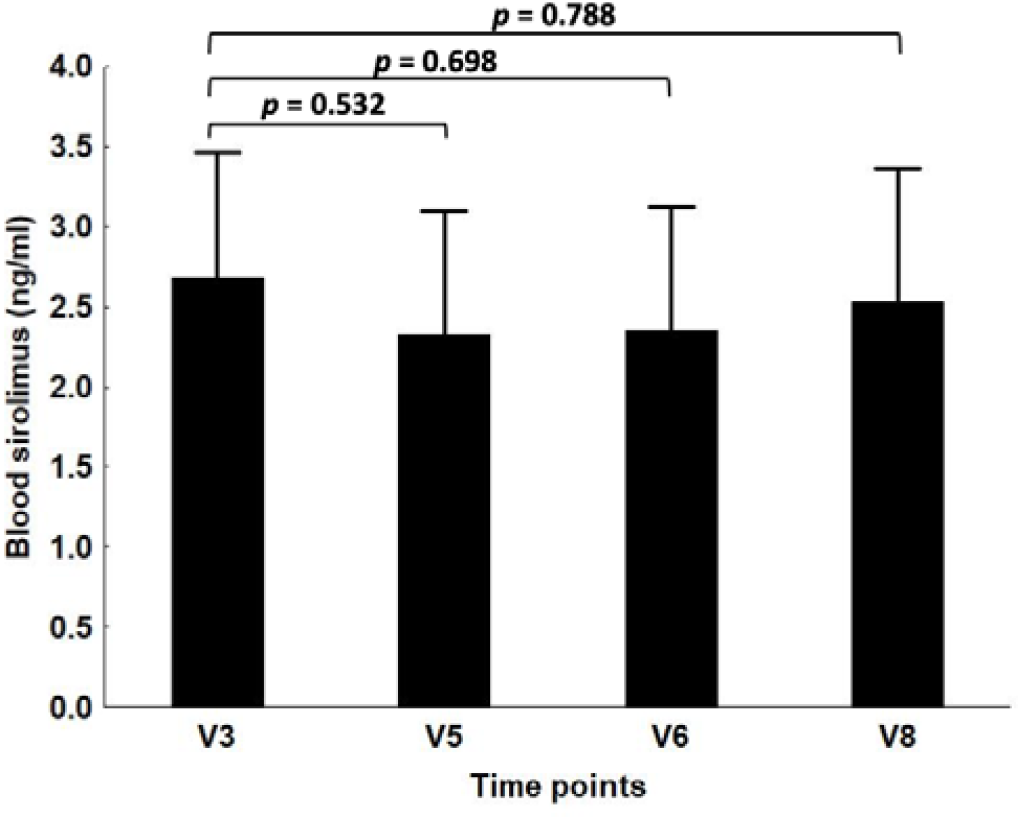
Blood levels of sirolimus (ng/ml). In calculating the average values, the samples with sirolimus blood concentrations <1.0 ng/ml were arbitrarily considered with a sirolimus concentration values = 1.0 ng/ml. When the *p* value comparing V8 to V3 was calculated, samples from patient n.24 were not considered.

### Evidence for increased γ-globin mRNA content in blood

In order to quantify the γ-globin mRNA content in the blood of sirolimus-treated patients we have developed an RT-qPCR technique performed to amplify mRNA sequences for γ globin and for RPL13A, GAPDH and β-actin control sequences starting from peripheral blood. This technique allowed to highlight an increase in γ-globin mRNA in the blood from most of the patients treated with sirolimus.

The experiment illustrated in **Figure 4(A-C)** demonstrates a clear increase in γ-globin mRNA in the blood of a patient treated with sirolimus and that this is not affected by the choice of the internal standards. In addition, the increase is certainly evident in V6.

**Figure 4.**
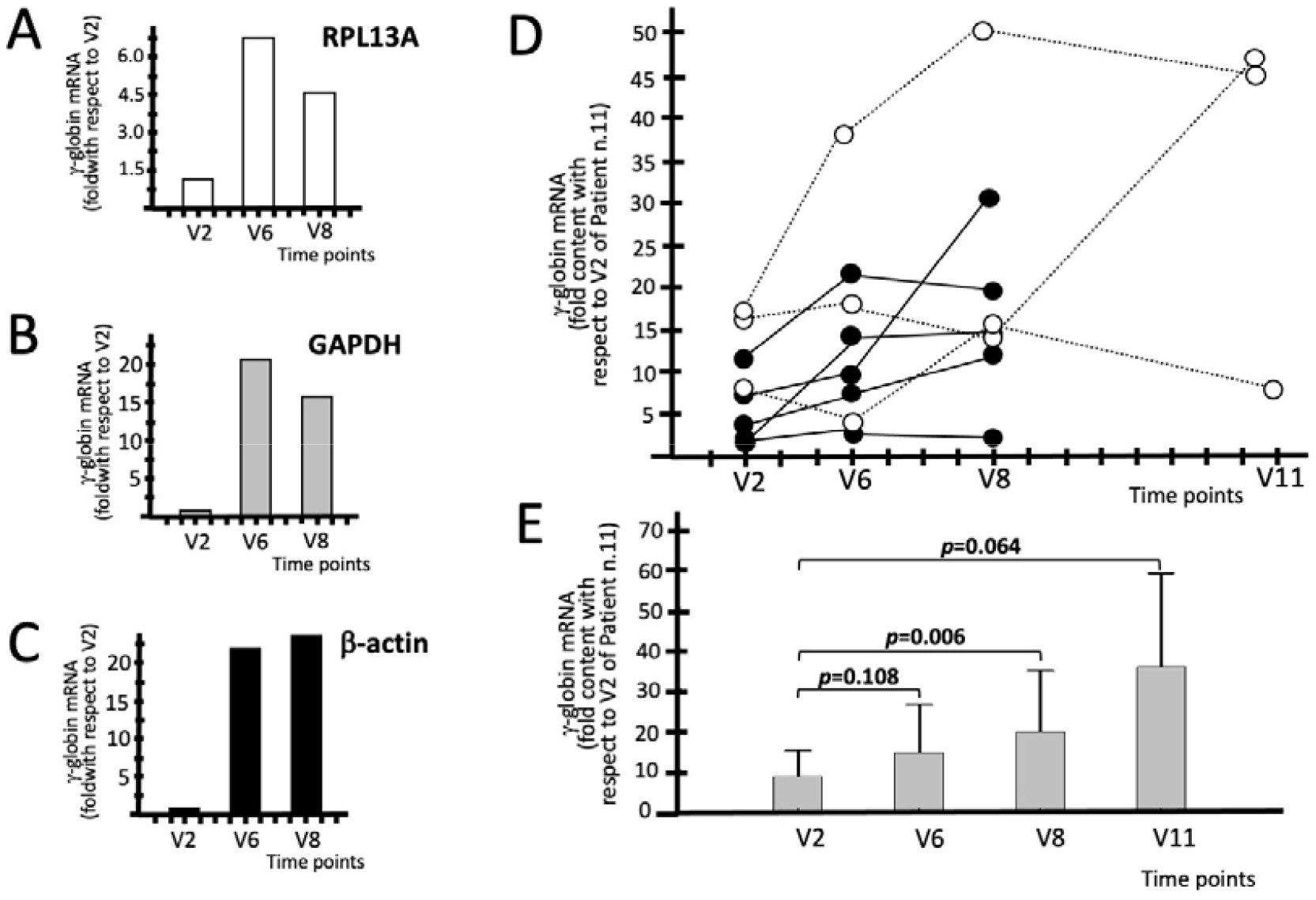
Increase of the content of γ-globin mRNA in the blood of patients treated with sirolimus. A-C. Representative data obtained with the blood samples of patient n.11 using RPL13A (A), GAPDH (B) and β-actin (C) control sequences, as indicated. D. Increase of the content of γ-globin mRNA in the blood of patients treated with sirolimus. The data represent the fold values with respect to V2 of patient n.11 (B). E. Average increase values of γ-globin mRNA expressed as fold content with respect to V2 of patient n.11 (E). Raw data are presented in **Supplementary Table S4**. N = 8 (V2-V8) and 3 (V11).

In analyzing the data obtained for all the sirolimus-treated patients we should consider that the patients are highly heterogeneous with respect to endogenous production of γ-globin mRNA. This is an expected result also considering the HPLC analyses of the patients recruited under the Sirthalaclin trial, as already reported in **Table III**. The % of HbF varies from 9.47% (patient n.21) to 21.32% (patient n.18).

The key results reported in **Figure 4D** demonstrate that increase of γ-globin mRNA content is clearly appreciable when the RT-qPCR data are normalized with respect to the V2 of patient n.11 (expressing the lower amounts of γ-globin mRNA). This calculation is important because gives information about the baseline levels of γ-globin mRNA present in the blood of analyzed patients. For instance, the blood samples of patient n.10 display a 17.71 relative value of γ-globin mRNA at V2 (in respect to the V2 value of patient n.11). Very interestingly, this value increased to 38.78 at V6, 50.83 at V8 and 48.8 at V11. Considering the late Sirthalaclin visits (V8 and V11) all the sirolimus-treated patients exhibited increased γ-globin mRNA content with the exception of patient n.21. As expected, when the data for each patient are compared to his/her V2 data, a similar trend was obtained. In this case the results are expressed as fold increase with respect to each V2. For instance, when the patient n.10 was considered, the γ-globin mRNA fold increase was 2.19 at V6, 2.87 at V8 and 2.76 at V11. **Figure 4E** shows the average increase of γ-globin mRNA in V6-V11, when all the data are reported relative to the V2 blood sample of patient n.11 (we performed this data analysis usually using as reference the sample exhibiting the lower amount of γ-globin mRNA, in this case patient n.11). The difference when V8 values are compared to V2 are statistically significant (*p* = 0.006) when the post-hoc test was performed. All the raw data are presented in **Supplementary Table S4**.

In conclusion, the RT-qPCR analysis using blood samples from sirolimus-treated patients show that the content of γ-globin mRNA increases in samples from all patients, with the exception of patient n.21.

### Evidence for increased γ-globin mRNA content in erythroid precursor cells (ErPCs)

Erythroid precursor cells have been isolated from sirolimus-treated patients at V6, V8 and V11. As it was done for the blood samples (see **Figure 4**, panel E and **Supplementary Table S4**), all the comparisons have been made with the V2 exhibiting the lower γ-globin mRNA level (in this case patient n.24).

The ErPC cultures were conducted in the presence of erythropoietin, but in the absence of sirolimus, in order to be sure that the differences observed are caused by the in vivo sirolimus treatment.

**Supplementary Table S5** and **Figure 5** report the data obtained by RT-qPCR, clearly demonstrating a borderline significant increase of γ-globin mRNA in ErPCs from sirolimus treated patients. This was observed both at V6 (*p* value = 0.060) and V8 (*p* value = 0.053). The trend was obtained in spite of the variability in the increase of γ-globin mRNA in V2 samples, reflecting baseline γ-globin mRNA levels in ErPCs of the different analyzed β-thalassemia patients. All the raw data are presented in **Supplementary Table S5**. It should be considered that sirolimus was absent during the erythropoietin treatment of the ErPCs; therefore, all the changes observed between the samples in V2 and the samples in V6, V8 and V11 should be ascribed to the in vivo effects of sirolimus on the studied patients.

**Figure 5.**
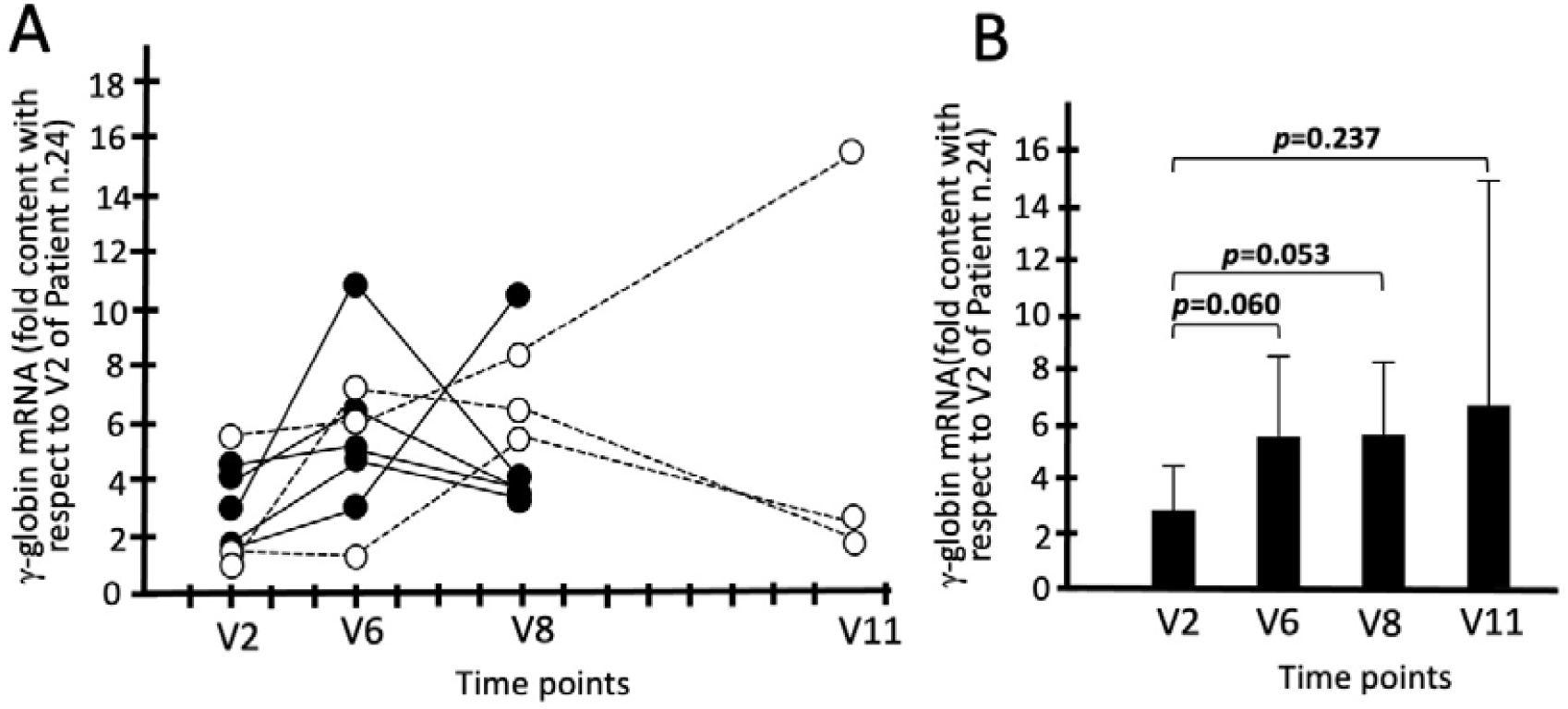
Increase of the content of γ-globin mRNA in the ErPCs isolated from patients treated with sirolimus. A. Increase of the content of γ-globin mRNA in the ErPCs of patients treated with sirolimus. The data represent the fold values with respect to V2 of patient n.24. B. Average increase of the content of γ-globin mRNA. The data represent the fold values with respect to V2 of patient n.24. Raw data are presented in **Supplementary Table S5**. N = 8 (V2-V8) and N = 3 (V11).

In conclusion, the RT-qPCR analysis using erythropoietin-cultured ErPC samples from sirolimus-treated patients support the concept that the expression of γ-globin genes increases in samples from all patients, with the exception of patients n.1 and n.21, that should be considered low-responders. In addition, it should be noted that the increase of sirolimus-induced γ-globin mRNA is already detectable at V6 both when samples of peripheral blood (see **Figure 4**) and of ErPCs (see **Figure 5)** are considered.

### Increase of HbF in erythroid precursor cells (ErPCs) from sirolimus-treated patients: HPLC analyses

**Figure 6A** shows representative HPLC pattern of V6 samples obtained stimulating with erythropoietin (EPO) ErPCs isolated from patients n.11 and n.18. In both cases, the % of HbF is increased in V6 samples compared with V2 samples. In patient n.18 samples the % of HbF increased from 16.33 (V2) to 29.38 (V6) (1.80 fold); in patient n.11 samples the % of HbF increased from 8.79 (V2) to 16.41 (V6) (1.88 fold). All the data obtained related to EPO-stimulated ErPCs from all the patients at V6 are shown in **Figure 6B**.

**Figure 6.**
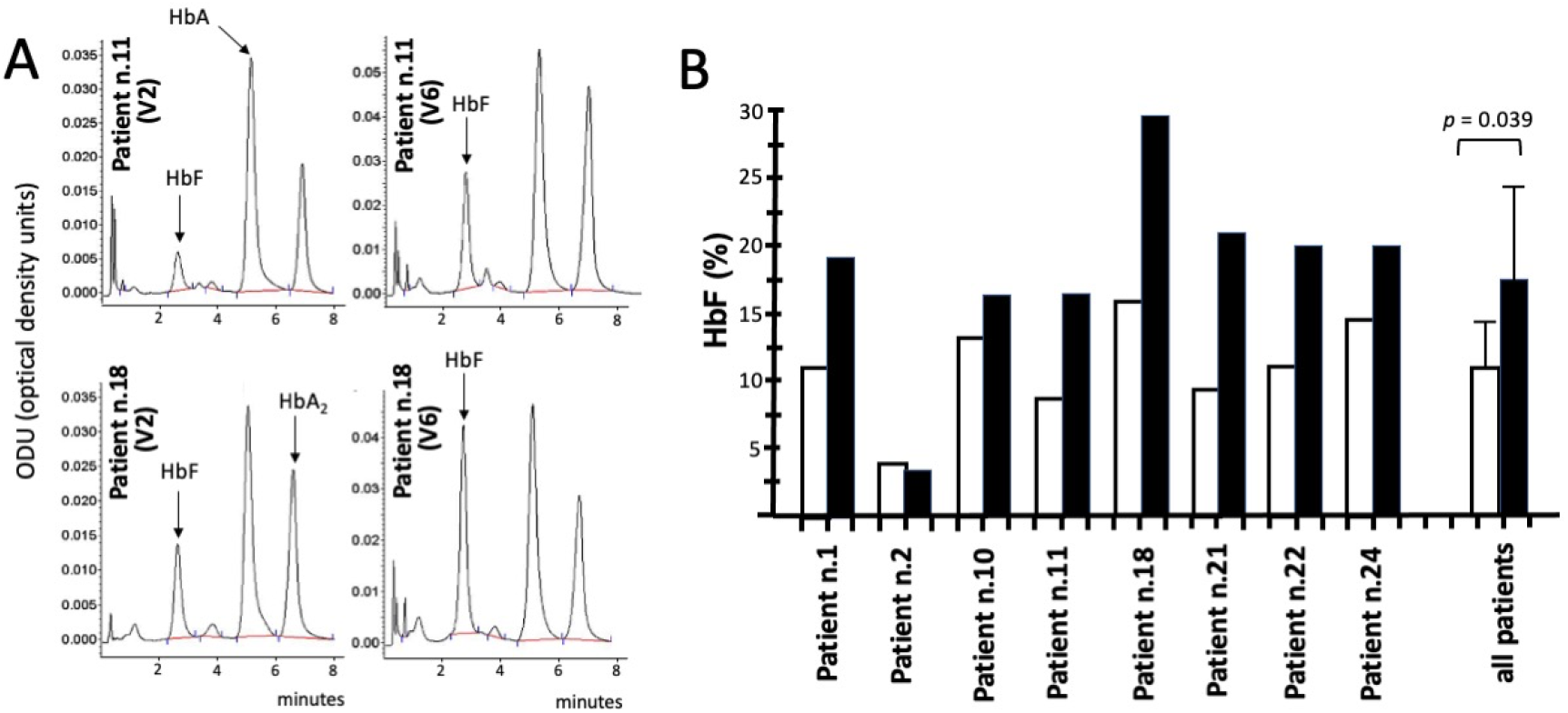
Increase of the % of HbF in EPO-induced ErPCs isolated at V6. A. Representative HPLC analysis of EPO-treated ErPCs from patients n.11 and n.18 at V2 and V6 Sirthalaclin visits. The arrows indicate the position of the HbA, HbA_2_ and HbF peaks. B. Changes of the % of HbF at V6 (black histograms) with respect to V2 (white histograms). The cumulative data from all the analyzed patients is also shown.

The data obtained, fully in agreement with the RT-qPCR data shown in **Supplementary Table S5** and **Figure 5A** demonstrate increased HbF production in ErPCs isolated at V6 from all the patients, with exception of patient n.2. The mean increase of HbF% was 1.57 that is statistically significant (*p* = 0.039).

In conclusion, the HPLC analysis using erythropoietin-cultured ErPC samples from sirolimus-treated patients strongly support the concept that HbF production is increased at V6, fully in agreement with the RT-qPCR data shown in **Figure 5**.

### Effects of sirolimus on biomarkers associated to ineffective erythropoiesis

Several biochemical markers are known to be associated to ineffective erythropoiesis and might be an important parameter to determine the activity in vivo of HbF inducers, including sirolimus [53-58]. The following parameters as index of erythroid response were measured before and under sirolimus treatment: erythroblast count, reticulocytes count and total bilirubin, LDH, soluble transferrin receptor (sTfR), erythropoietin and ferritin levels. While no significant difference under treatment with sirolimus versus baseline in erythroblast count, reticulocytes count, LDH and erythropoietin levels was observed (data not shown), a significant reduction was found in total bilirubin levels, sTfR, and ferritin levels. In addition to these markers, we have analyzed the possible inhibitory effects of sirolimus on the excess of free α-globin chains produced by ErPCs. In this context, the excess of free α-globin chains has been firmly demonstrated to correlate with red-blood toxicity. It is known that all these parameters participate to a complex network affecting erythropoiesis (and ineffective erythropoiesis) and iron metabolism balance [54-58].

**Figure 7** shows the sirolimus-mediated changes of free α-globin chains, bilirubin, sTfR and ferritin. The data obtained indicate that there is a consistent reduction of free α-globin chains in ErPCs isolated from β-thalassemia patients treated with sirolimus (**Figure 7A**). This reduction was found to be significant at V6 (*p* = 0.007) and V8 (*p* = 0.002) time periods. This is relevant, since the excess of free α-globin chains is one of the most important causes of red blood cell toxicity.

**Figure 7.**
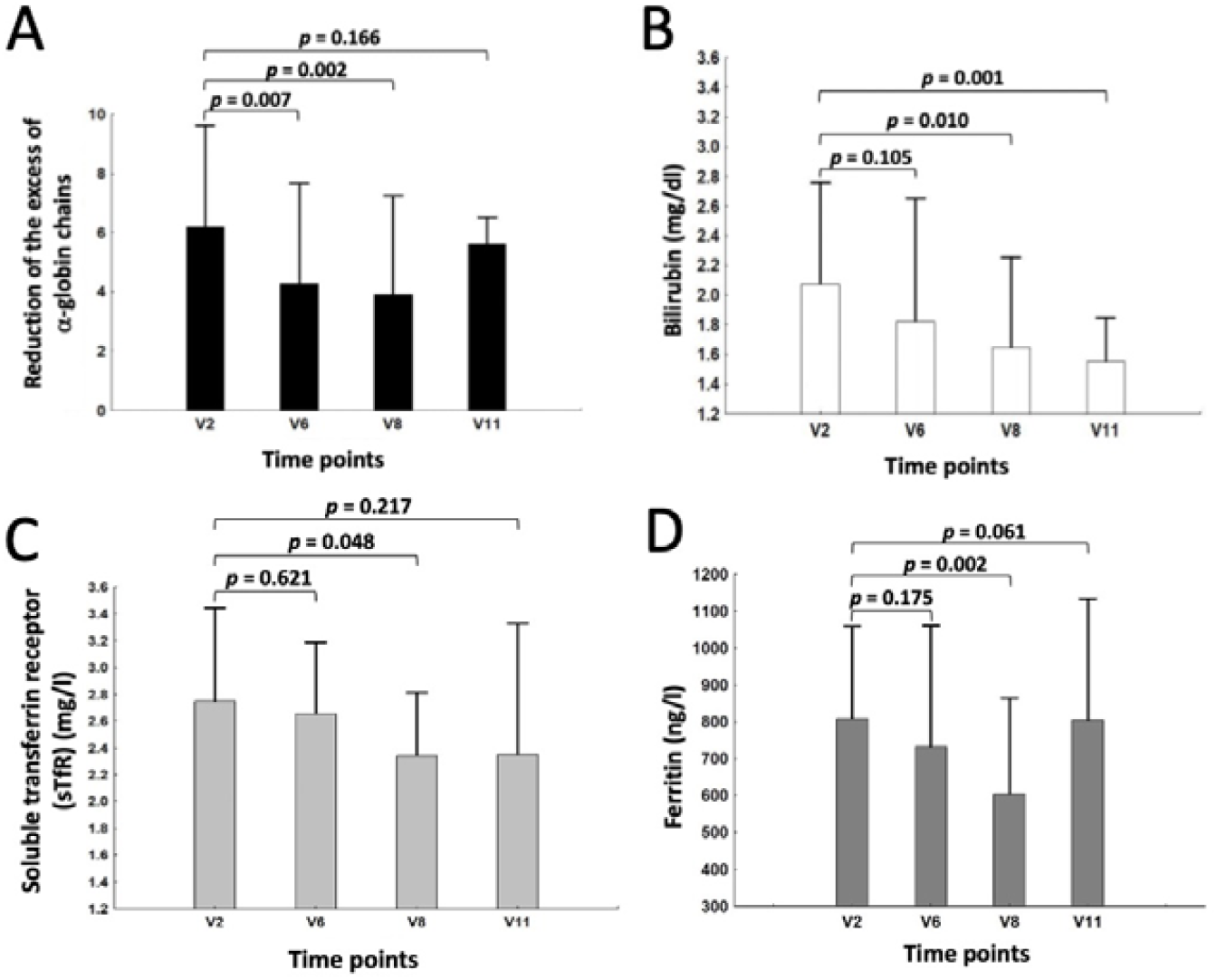
Changes of free α-globin chains (A), bilirubin (B), soluble transferrin receptor (C) and ferritin (D) following treatment with sirolimus. Only six patients were analyzed for changes of free α-globin chains, since two patients exhibited lack of detectable α-globin peak after HPLC analysis. Raw data related to panels B-D are presented in **Supplementary Tables S6, S7 and S8**. N = 8 (V2-V8) and 3 (V11).

Moreover, a statistically significant decrease at V8 of bilirubin (**Figure 7B**), sTfR (**Figure 7C**) and ferritin (**Figure 7D**) was also found. All these are markers of ineffective erythropoiesis and have been demonstrated to be expressed at high levels in the plasma of β-thalassemia patients. Notably, the significance of these changes was found at V8 and V11, for bilirubin, time periods, i.e. after the increase of γ-globin and HbF found at V6 (see **Figure 5** and **Figure 6**). Overall, the data suggest some positive impact of the sirolimus treatment on ineffective erythropoiesis.

### Evidence for a clinical effect in sirolimus-treated patients at the end of the treatment: indexes of transfusion demand and blood HbF

The index of transfusion demand was calculated dividing the red cell consumption by the average pre-transfusion hemoglobin concentration (see Materials and methods section). If the endogenous production of hemoglobin was increases under sirolimus treatment, then this parameter was expected to decrease proportionally. As shown in **Figure 8** and **Table V**, a reduction of this parameter was interestingly observed. This reduction was found statistically significant (*p* = 0.006) (**Figure 8**).

**Figure 8.**
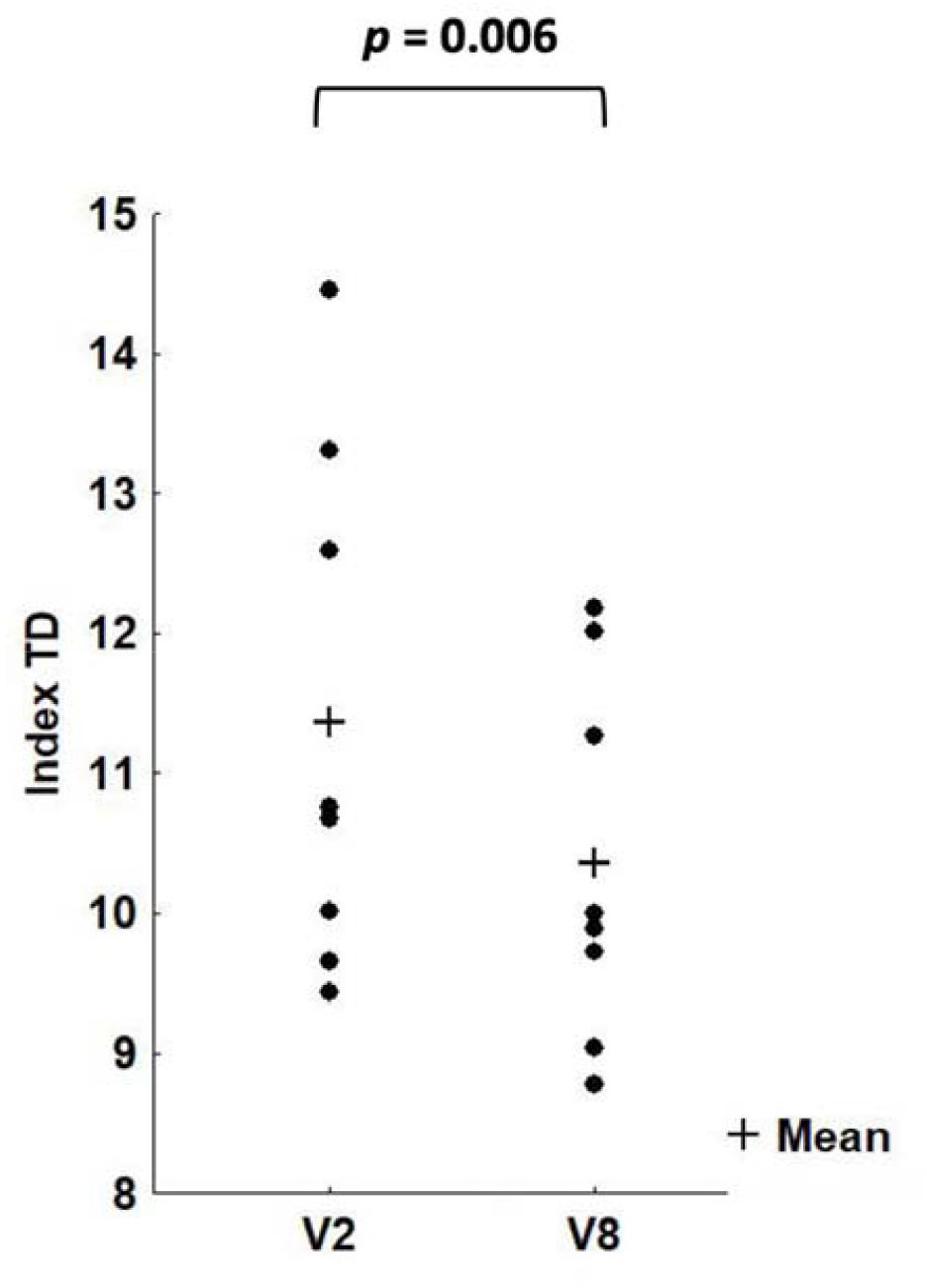
Changes in the transfusion demand (TD) when data at V2 and V8 are considered (*p* = 0.006). N = 8.

**TABLE V.**
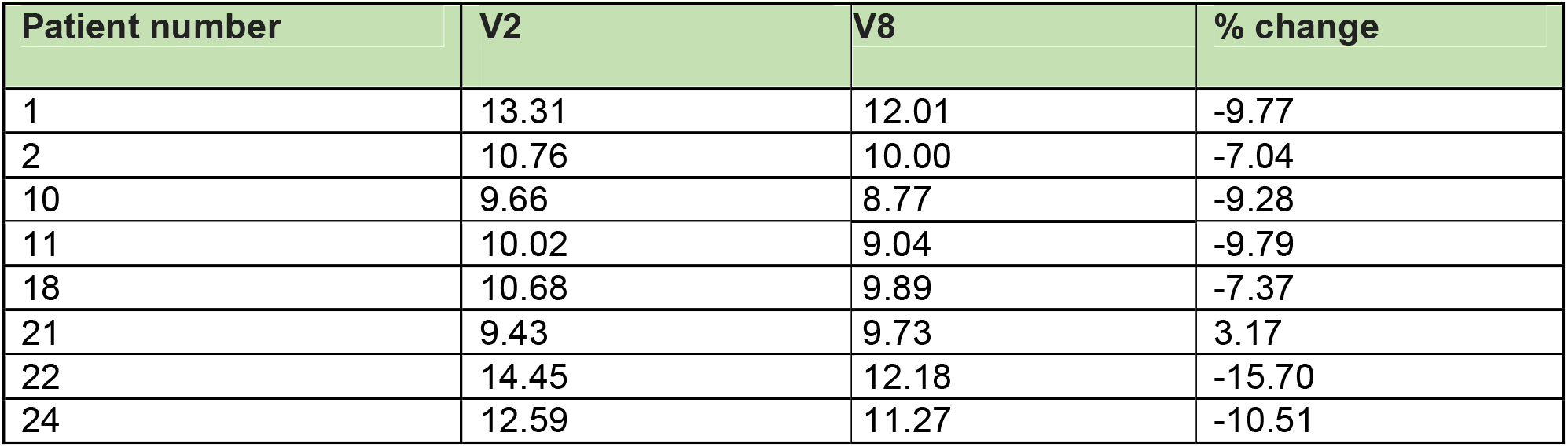
Index of transfusion demand (ml/kg/year/gHb)

In parallel, an increase of blood HbF was found at the end of the sirolimus treatment in all the patients with the exception of patients n.21 and n.22 (the % change with respect to V2 was -51% and -23%, respectively). The average fold increase of HbF was 1.22 (S.D. 0.33) and close to statistical significance (*p* = 0.076). If the patient n.21 (not responder also in the transfusion demand assay) was not considered, the new calculated fold increase was highly significant (p < 0.001). The best increase of blood HbF was found in the blood of patients n.1, n.2 and n.18 (+38%, +49% and +55%, respectively). Intermediate HbF blood increase (+21/+22%) was found in patients n.10, n.11 and n.24.

Notably, the parameter of patient n.21 (increase of transfusion demand and no increase of HbF content) were in agreement with the lack of increase in the content of γ-globin mRNA in the blood.

### Immunophenotype before and after sirolimus administration

As shown in **Figure 9**, the patients did not show significant changes in the pattern of lymphocyte parameters. The analysis was performed by flow cytometric technique (FACS) analyzing the following markers associated with the various lymphocyte subpopulations: CD3, CD4, CD8, CD14, CD19, CD25. The starting material for the analysis consisted of PBMCs obtained from the peripheral blood of the patients. The PBMCs extracted from the blood were then frozen and stored in liquid nitrogen until the time of analysis. Before proceeding with the incubation with specific antibodies to detect the aforementioned markers, a molecule (Aqua Dead Cell Stain) was also used, capable of discriminating between living and dead cells, in order to accurately analyze only the intact cells after defrosting. As an example, we show the data obtained analyzing the cells from patient n.18 with the FACS apparatus (**Figure 9A**).

**Figure 9.**
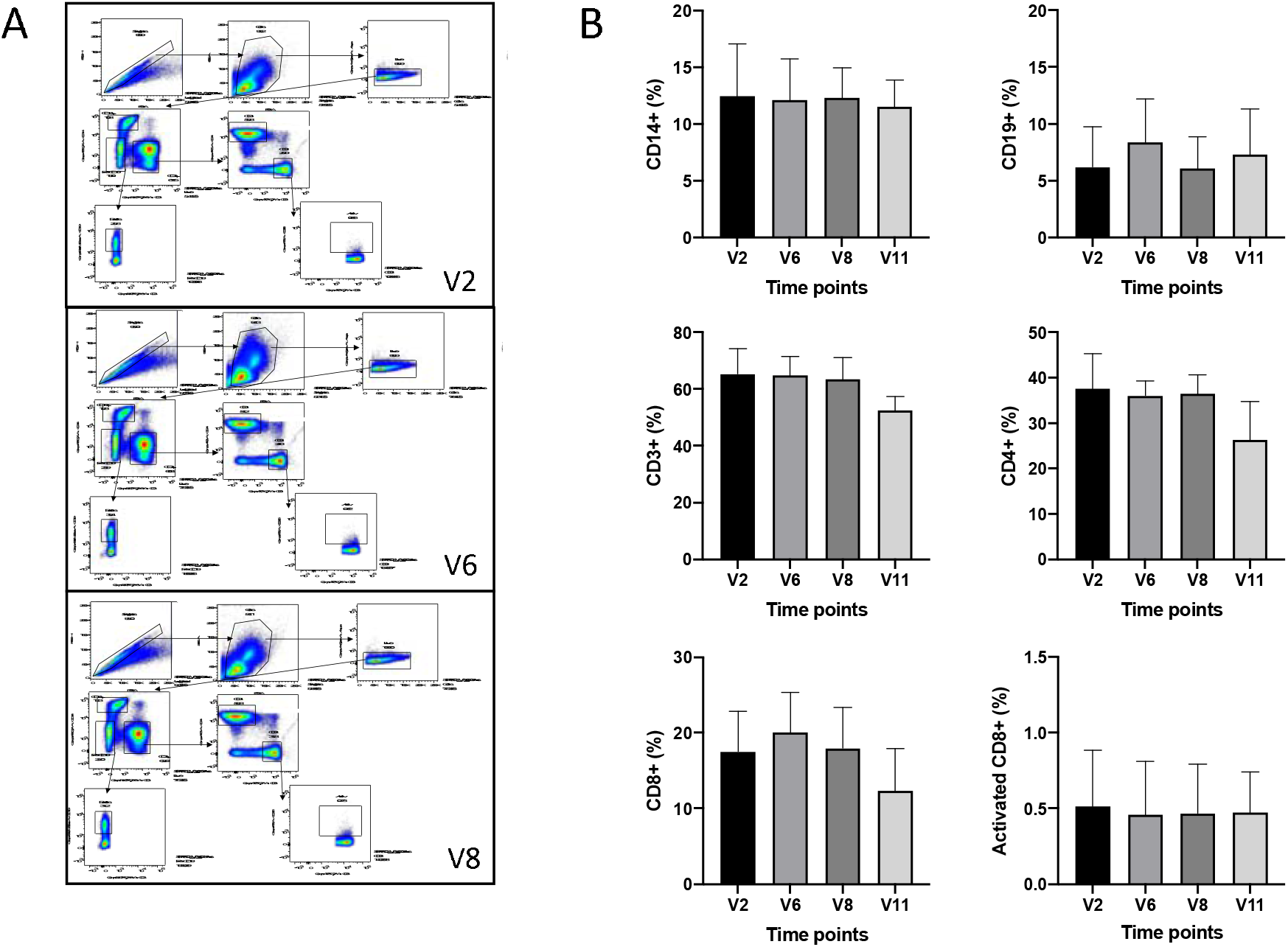
A. Representative example (samples from patient n.18) of FACS analysis showing the employed gating strategy with the following markers associated with the various lymphocyte subpopulations: CD3, CD4, CD8, CD14, CD19, CD25. B. Average changes of the immunophenotype pattern at V2, V6, V8 and V11. N = 8 (V2-V8) and 3 (V11).

As can be seen from **Figure 9B**, the patients analyzed did not undergo significant percentage variations in the various cellular subsets obtained by immunophenotyping (p > 0.05) with the exception of V11 samples related to CD3+ and CD4+, which resulted significantly lower (p <0.031 and p <0.022, respectively). Therefore, we can conclude that even after 12 months of chronic therapy with sirolimus, thalassemia patients show only minor alterations in the profile in lymphocyte typing. The recorded values are expressed as a percentage of positive cells for a given marker with respect to the totality of live cells analyzed, with the exception of effector T lymphocytes (activated CD8+) which are expressed as a percentage of CD8+ lymphocytes.

### Adverse Effects (AE)

Adverse Effects AEs were classified as expected or un-expected events on their severity (serious, mild, moderate, severe) according to the Common Terminology Criteria for Adverse Events (Version 5.0). They were also considered as related or not to the investigational drug. No major adverse effects were found in the treated β-thalassemia patients. However, despite detailed instructions given to each patient in order to prevent and to early treat oral lesions, 11 episodes of stomatitis, consistent with minor side effects associated with the activity of mTOR inhibitors [59-62] were observed in 5 patients and classified as mild and moderate. Three patients independently decided to interrupt the trial due to this recurrence of stomatitis. A detailed analysis of the observed stomatitis is reported in **Supplementary Table S9**.

## Discussion

SIRTHALACLIN was the first clinical trial in β-thalassemia based on the use of sirolimus [41-43]. This open label monocentric study was planned to enroll about 30 transfusion dependent patients with β^+^/β^+^ and β^+^/β^0^ thalassemia genotype in order to treat with oral sirolimus 20 patients whose Erythroid Precursor Cells (ErPCs) were responsive in vitro to sirolimus (level of fetal hemoglobin, sirolimus concentration 100 nanomolar). Total enrollment (signature of the informed consent) was 24 patients and in vitro response was present in 17, however only 8 decided to take sirolimus by the oral route (0.5 to 2.0 mg daily). None of the patients had to stop sirolimus for safety reason. Out of the 8 patients we have data for all of them at 6 months and for three of them at 12 months.

The main purpose of this study was to determine whether treatment with sirolimus induces an increase of the expression of γ-globin genes in treated β-thalassemia patients. Several studies were published supporting this trial. First, sirolimus was found to induce HbF accumulation in erythroid precursors isolated from β-thalassemia and sickle-cell disease patients [28-31]. Second, in vivo experiments using animal systems have demonstrated that sirolimus treatment increases red blood cell counts and hemoglobin levels, supporting the concept that sirolimus administration may be of help in patients with ineffective erythropoiesis [53]. In addition, sirolimus was found to induce γ-globin genes, accumulate HbF and improve anemia in model systems of in sickle cell disease (SCD) [33]. Sirolimus treatment prolonged the lifespan of sickle cell erythrocytes in circulation, reduced spleen size, and reduced renal and hepatic iron accumulation in SCD mice [34]. Third, sirolimus or sirolimus analogues (either alone or in combination with hydroxycarbamide) induced HbF production in patients treated who underwent kidney transplantation [35,36]. These studies demonstrated that in SCD patients treated with sirolimus or sirolimus analogues the HbF level was dramatically increased.

In addition to the effects on γ-globin genes and HbF production, sirolimus has been demonstrated to retain other biological activities that are expected to be useful in the treatment of hemoglobinopathies, including β-thalassemia. In a mouse model of thalassemia, Lechauve et al. [53] found that treatment with sirolimus was associated with a significant reduction in α-globin accumulation and ineffective erythropoiesis along with a longer life span for red blood cells, a combination of effects potentially important for patients [53]. Another possible in vivo effect of sirolimus might be an enhancement of Hematopoietic Stem Cells (HSCs) through regulation of the mTOR pathway [63]. This interesting possibility is sustained by findings suggesting that sirolimus, together with other biological response modifiers, participates in vivo in increasing the number of long-term HSCs [58].

Information on this study (focusing on β^+^/β^+^ and β^+^/β^0^ thalassemia genotypes) can be found at NCT03877809 [41] and in the review paper by Gamberini et al. [42]. In addition, a second study is ongoing (NCT04247750) focusing on β^0^/β^0^ and β^+^/β^0^ thalassemia genotypes [42].

The first finding of the study here presented is that the expression of γ-globin mRNA is increased in blood and erythroid precursor cells isolated from β-thalassemia patients treated with low-dose sirolimus. This effect was found early during the trial (i.e. after 90 days treatment, at the V6 visit). This finding was fully in agreement with the HbF accumulation by erythroid precursors (ErPCs) from sirolimus-treated β-thalassemia patients. When HPLC was performed at V6 an increase of HbF was found. It should be underlined that ErPCs isolated from sirolimus-treated β-thalassemia patients have been cultured with only SCF and EPO (without sirolimus), therefore suggesting that the changes in gene expression found should be ascribed to the in vivo treatment with sirolimus. This is the first description of an increase of γ-globin mRNA expression in β-thalassemia patients treated with low-dose sirolimus.

The interest on sirolimus is related to the fact that sirolimus is a well-known agent, used for many years for other indications [64-77] (see **Supplementary Table S10**). For instance, sirolimus or sirolimus-related compounds have been employed for treatment of kidney transplantation [64,65], cardiac [66] and liver [67] transplantation, lupus erythematosus (SLE) [68], autoimmune cytopenias [69], lymphangioleiomyomatosis (LAM) [70], tuberous sclerosis complex [71], recurrent meningioma [72], pancreatic neuroendocrine tumors (NET) [73], advanced differentiated thyroid cancers [74], advanced breast cancer [75], B-cell lymphomas [76], metastatic renal cell carcinoma [77].

In respect to clinical management of β-thalassemia patients, sirolimus was considered a repurposed drug when the first evidence of its ability to induce in vitro HbF production in erythroid precursor cells from β-thalassemia and sickle cell disease patients was reported and further validated [28-31].

A second important conclusion of our trial is that sirolimus has effects on erythropoiesis and reduces biochemical markers associated to ineffective erythropoiesis. For instance, ErPCs from sirolimus-treated patients exhibited a clear reduction of the excess of free α-globin chains. The targeting of the network(s) regulating this parameter might be very important, since in β-thalassemia, accumulated free α-globin forms intracellular precipitates that impair erythroid cell maturation and viability. Induction of autophagy is expected to help in activating quality control systems able to mitigate β-thalassemia pathophysiology by degrading toxic free α-globin [53,78,79]. The effects of on autophagy-associated parameters will be the object of a successive study.

In addition to the reduction of accumulating free α-globin, sirolimus treatment was able to modify some other biochemical parameters associated to ineffective erythropoiesis, such as bilirubin, soluble transferrin receptor and ferritin. All these parameters are higher in β-thalassemia patients with respect to unaffected population. We can speculate that the change in ferritin levels could be regarded as an indication of the modification of iron metabolism in its complex network with erythropoietic activity [80,81]. The sirolimus-mediated reduction of these biochemical parameters might indicate an improvement in ineffective erythropoiesis. Moreover, these effects of sirolimus appeared late in the treatment (starting from V8).

These data suggest that additional effects of sirolimus in vivo might occur, in addition to the stimulation of γ-globin gene expression and HbF production.

As far as clinical data, transfusion burden was evaluated as index of transfusion demand, since it is adjusted for the pre-transfusion hemoglobin concentration. No patient showed a major reduction in transfusion burden, but in the sirolimus-treated cohort the majority of the patients (5 out of 8 patients) displayed a significant reduction of the index of transfusion demand (*p* = 0.0056) in the first six months of therapy.

In order to comment on this, we would like to remind that this trial was designed to verify whether positive effects of sirolimus would be obtained in β-thalassemia patients without altering their transfusion regimen and using low-dosages of sirolimus.

As far as the first point, it would be very interesting in testing sirolimus on NTDT (non transfusion-dependent thalassemia) as well as on β-thalassemia patients not regularly receiving blood transfusions. On the other hand, the data presented in the present paper should be in the future considered together the results that will be obtained during the trial NCT04247750, focused on patients carrying with β^0^/β^0^ and β^+^/β^0^ thalassemia genotypes.

As far as the second point, we have to point out that the sirolimus content in the blood was found to be variable and low. With regard to variability, the values observed cannot be considered unexpected, since high variability has been found in clinical trials carried out in other diseases [50-52]. With regard to overall blood content, it was found lower than expected from published data on other pathologies (**Figure 3** and **Table IV**) [50-52]. For instance, in 90 patients with lymphangioleiomyomatosis the average blood sirolimus level was 7.2□±□2.6□ng/ml (range 1.5 to 18.6□ng/ml) in the first year of sirolimus treatment under the average dose of 1.27□±□0.47□mg/day (range 0.5 to 2□mg/day) [51]. In our patients the actual average dose was 1.2 mg/day at 3 months and 1.7 mg/day at 6 months. This sirolimus dosage was associated with a blood concentration of 2.2 ± 0 .9 ng/ml after 3 months of treatment (range 1.1 to 4.5) and 3.0 ± 1.6 ng/ml after 6 months of treatment (range 1.0 to 4.6). When these data are compared with the HPLC and RT-qPCR data, it should be concluded that these low levels of sirolimus are sufficient to cause changes in γ-globin gene expression.

Under these circumstances the drug was well tolerated without alteration of the immunophenotype. While no major side effects were observed, five patients exhibited stomatitis (for three of them this effect was recurrent). Our study demonstrates that this was the only relevant side effect of the treatment, as reported also in other cohorts of patients affected by other pathologies (TubulusSchlerosis Complex, lymphangioleiomyomatosis, organ transplantation, cancer), treated with mTOR inhibitors [59]. This issue should be considered in the case other sirolimus-based clinical interventions are proposed. In this context, good oral hygiene should be maintained [60]. and spicy, acidic, hot, or hard foods avoided. In addition, topical high-potency corticosteroids (e.g.: clobetasol, dexamethasone), nonsteroidal anti-inflammatory drugs (NSAIDs; eg, amlexanox paste), and anesthetics (e.g.: viscous lidocaine) can be considered. The issues related to possible side effects of mTOR inhibitors are reviewed more extensively elsewhere [59-62]. Alternatively, future experimental trials might consider the use of other rapalogs.

In conclusion, despite the limited number of patients and the variability of response, the data indicates that sirolimus given at low doses modifies hematopoiesis and induces increased expression of γ-globin genes in a sub-set of β-thalassemia patients. Further clinical trials are warranted, considering the possibility to test the drug in patients with less severe forms of the disease, exploring combination therapies and other end points not considered in the present trial.

## Data Availability

All of the data produced in the present work are contained in the manuscript and in the supplementary material. Other information related to the present study is available upon reasonable request to the authors.

## Acknowledgements

The authors wish to thank all the patients involved in the trial. The activity of the CRO was greatly appreciated. C.Z., L.C.C., M.Z. and C.P. have been supported by a fellowship from “Tutti per Chiara Onlus”. This paper is dedicated to the memory of Chiara Gemmo and Elio Zago. We thank ALT (Associazione per la lotta alla Talassemia “Rino Vullo”-Ferrara), AVLT (Associazione Veneta per la Lotta alla Talassemia “Elio Zago” -APS), and Interuniversity Consortium for the Biotechnology (C.I.B., Italy) for support. We thank Dr. Raffaella Vergura (Project Manager, MTA, Medical Trials Analysis, Ferrara, Italy) for the organization and management of the trial.

## Abbreviations

Hb: hemoglobin
HbA: adult hemoglobin
HbF: fetal hemoglobin
HPFH: hereditary persistence of fetal hemoglobin
TDT: transfusion-dependent thalassemia
SCD: sickle-cell disease
HU: hydroxyurea
mTOR: mammalian target of rapamycin
ROS: reactive oxygen species
hHSPCs: Human Hematopoietic Stem and Progenitor Cells
ErPCs: erythroid precursor cells
sTFR: soluble transferrin receptor
SLE: systemic lupus erythematosus
LAM: lymphangioleiomyomatosis
HIV: human immunodeficiency virus
HBV: hepatitis B virus
HCV: hepatitis C virus
FACS: fluorescence-activated cell sorter
NGS: next-generation sequencing
AEs: adverse effects
EU: European Union
ODD: orphan drug designation
EMA: European Medicinal Agency
FDA: Food and Drug Administration

## References

1. Weatherall DJ. Phenotype-genotype relationships in monogenic disease: lessons from the thalassaemias. Nat Rev Genet 2001; 2: 245–55.

2. Origa R. β-Thalassemia. Genet Med 2017; 19: 609–619.

3. Fucharoen S, Weatherall DJ. Progress Toward the Control and Management of the Thalassemias. Hematol Oncol Clin North Am 2016; 30: 359–71.

4. Modell B, Darlison M, Birgens H, et al. Epidemiology of haemoglobin disorders in Europe: an overview. Scand J Clin Lab Invest 2007; 67: 39–69.

5. Galanello R, Origa R. B-thalassemia. Orphanet J Rare Dis 2010; 5: 11.

6. Colah R, Gorakshakar A, Nadkarni A. Global burden, distribution and prevention of β-thalassemias and hemoglobin E disorders. Expert Rev Hematol 2010; 3: 103–17.

7. Modell B, Darlison M. Global epidemiology of haemoglobin disorders and derived service indicators. Bull World Health Organ 2008; 86: 480–7.

8. Thein SL. The molecular basis of β-thalassemia. Cold Spring Harb Perspect Med 2013; 3: a011700

9. Sripichai O, Fucharoen S. Fetal hemoglobin regulation in β-thalassemia: heterogeneity, modifiers and therapeutic approaches. Expert Rev Hematol 2016; 9: 1129–1137.

10. Liu D, Zhang X, Yu L, et al. KLF1 mutations are relatively more common in a thalassemia endemic region and ameliorate the severity of β-thalassemia. Blood 2014; 124: 803–811

11. Musallam KM, Sankaran VG, Cappellini MD, et al. Fetal hemoglobin levels and morbidity in untransfused patients with β-thalassemia intermedia. Blood 2012; 119: 364–367.

12. Forget BG. Molecular basis of hereditary persistence of fetal hemoglobin. Ann N Y Acad Sci 1998; 850: 38–44.

13. Nuinoon M, Makarasara W, Mushiroda T, et al. A genome-wide association identified the common genetic variants influence disease severity in β0-thalassemia/hemoglobin E. Hum Genet 2010; 127: 303–314.

14. Galanello R, Sanna S, Perseu L, et al. Amelioration of Sardinian β0 thalassemia by genetic modifiers. Blood 2009; 114: 3935–3937.

15. Danjou F, Anni F, Perseu L, et al. Genetic modifiers of β-thalassemia and clinical severity as assessed by age at first transfusion. Haematologica 2012; 97: 989–993.

16. Badens C, Joly P, Agouti I, et al. Variants in genetic modifiers of β-thalassemia can help to predict the major or intermedia type of the disease. Haematologica 2011; 96: 1712–1714.

17. Uda M, Galanello R, Sanna S, et al. Genome wide association study shows BCL11A associated with persistent fetal hemoglobin and amelioration of the phenotype of β-thalassemia. Proc Natl Acad Sci USA 2008; 105: 1620–1625.

18. Gambari R, Fibach E. Medicinal chemistry of fetal hemoglobin inducers for treatment of β-thalassemia. Curr Med Chem 2007; 14: 199–212.

19. Lavelle D, Engel JD, Saunthararajah Y. Fetal Hemoglobin Induction by Epigenetic Drugs. Semin Hematol 2018; 55: 60–67.

20. Finotti A, Gambari R. Recent trends for novel options in experimental biological therapy of β-thalassemia. Expert Opin Biol Ther 2014; 14: 1443–54.

21. Nuamsee K, Chuprajob T, Pabuprapap W, et al. Trienone analogs of curcuminoids induce fetal hemoglobin synthesis via demethylation at (G)gamma-globin gene promoter. Sci Rep 2021; 11: 8552.

22. Antoniani C, Meneghini V, Lattanzi A, et al. Induction of fetal hemoglobin synthesis by CRISPR/Cas9-mediated editing of the human β-globin locus. Blood 2018; 131: 1960–1973.

23. Métais JY, Doerfler PA, Mayuranathan T, et al. Genome editing of HBG1 and HBG2 to induce fetal hemoglobin. Blood Adv 2019; 3: 3379–3392.

24. Mingoia M, Caria CA, Ye L, et al. Induction of therapeutic levels of HbF in genome-edited primary β(0) 39-thalassaemia haematopoietic stem and progenitor cells. Br J Haematol 2021; 192: 395–404.

25. Frangoul H, Altshuler D, Cappellini MD, et al. CRISPR-Cas9 Gene Editing for Sickle Cell Disease and β-Thalassemia. N Engl J Med 2021; 384: 252–260.

26. NCT03655678 (A Safety and Efficacy Study Evaluating CTX001 in Subjects With Transfusion-Dependent β-Thalassemia)

27. Sehgal SN. Sirolimus: its discovery, biological properties, and mechanism of action. Transplant Proc 2003; 35(3 Suppl): 7S–14S.

28. Mischiati C, Sereni A, Lampronti I, et al. Rapamycin-mediated induction of gamma-globin mRNA accumulation in human erythroid cells. Br J Haematol 2004; 126: 612–21.

29. Fibach E, Bianchi N, Borgatti M, et al. Effects of rapamycin on accumulation of alpha-, β- and gamma-globin mRNAs in erythroid precursor cells from β-thalassaemia patients. Eur J Haematol 2006; 77: 437–41.

30. Zuccato C, Bianchi N, Borgatti M, et al. Everolimus is a potent inducer of erythroid differentiation and gamma-globin gene expression in human erythroid cells. Acta Haematol 2007; 117: 168–76.

31. Pecoraro A, Troia A, Calzolari R, et al. Efficacy of Rapamycin as Inducer of Hb F in Primary Erythroid Cultures from Sickle Cell Disease and β-Thalassemia Patients. Hemoglobin 2015; 39: 225–9.

32. Zhang X, Campreciós G, Rimmelé P, et al. FOXO3-mTOR metabolic cooperation in the regulation of erythroid cell maturation and homeostasis. Am J Hematol 2014; 89 :954–63.

33. Khaibullina A, Almeida LE, Wang L, et al. Rapamycin increases fetal hemoglobin and ameliorates the nociception phenotype in sickle cell mice. Blood Cells Mol Dis 2015; 55: 363–72.

34. Wang J, Tran J, Wang H, et al. mTOR Inhibition improves anaemia and reduces organ damage in a murine model of sickle cell disease. Br J Haematol 2016; 174: 461–9.

35. Gaudre N, Cougoul P, Bartolucci P, et al. Improved Fetal Hemoglobin With mTOR Inhibitor-Based Immunosuppression in a Kidney Transplant Recipient With Sickle Cell Disease. Am J Transplant 2017; 17: 2212–2214.

36. Al-Khatti AA, Alkhunaizi AM. Additive effect of sirolimus and hydroxycarbamide on fetal haemoglobin level in kidney transplant patients with sickle cell disease. Br J Haematol 2019; 185: 959–961.

37. Ansari SH, Lassi ZS, Khowaja SM, et al. Hydroxyurea (hydroxycarbamide) for transfusion-dependent β-thalassaemia. Cochrane Database Syst Rev 2019; 3: CD012064.

38. Algiraigri AH, Wright NAM, Paolucci EO, Kassam A. Hydroxyurea for lifelong transfusion-dependent β-thalassemia: A meta-analysis. Pediatr Hematol Oncol 2017; 34: 435–448.

39. Foong WC, Ho JJ, Loh CK, Viprakasit V. Hydroxyurea for reducing blood transfusion in non-transfusion dependent β thalassaemias. Cochrane Database Syst Rev 2016; 10: CD011579.

40. Paolo Rigano, Alice Pecoraro, Roberta Calzolari, et al. Desensitization to hydroxycarbamide following long-term treatment of thalassaemia intermedia as observed in vivo and in primary erythroid cultures from treated patients. Br J Haematol 2010; 151: 509–15.

41. NCT03877809 (A Personalized Medicine Approach for β-thalassemia Transfusion Dependent Patients: Testing sirolimus in a First Pilot Clinical Trial: SIRTHALACLIN)

42. Gamberini MR, Prosdocimi M, Gambari R. Sirolimus for Treatment of β-Thalassemia: From Pre-Clinical Studies to the Design of Clinical Trials. Health Education and Public Health 2021; 4: 425–435.

43. Musallam KM, Bou-Fakhredin R, Cappellini MD, Taher AT. 2021 update on clinical trials in beta-thalassemia. Am J Hematol 2021.

44. Cappellini MD., Farmakis D., Porter J., et al. Guidelines for the Management of Transfusion Dependent Thalassaemia (4th Edition – 2021). Thalassemia International Federation, 2021.

45. Wang M, Tang DC, Liu W, et al. Hydroxyurea exerts bi-modal dose-dependent effects on erythropoiesis in human cultured erythroid cells via distinct pathways. Br J Haematol 2002;119: 1098–105.

46. Fibach E, Bianchi N, Borgatti M, et al. Mithramycin induces fetal hemoglobin production in normal and thalassemic human erythroid precursor cells. Blood 2003; 102: 1276–81.

47. Verschoor, C. P., Kohli, V., Balion, C. A Comprehensive Assessment of Immunophenotyping Performed in Cryopreserved Peripheral Whole Blood. Cytometry Part B 2018; 94B: 818–826.

48. Finak, G., Langweiler, M., Jaimes, M., et al. Standardizing Flow Cytometry Immunophenotyping Analysis from the Human ImmunoPhenotyping Consortium. Sci Rep 2016; 6: 20686.

49. Pinto, L.A., Trivett, M.T., Wallace, D., et al. Fixation and cryopreservation of whole blood and isolated mononuclear cells: Influence of different procedures on lymphocyte subset analysis by flow cytometry. Cytometry 2005; 63B: 47–55.

50. Lai NS, Yu HC, Huang KY, et al. Decreased T cell expression of H/ACA box small nucleolar RNA 12 promotes lupus pathogenesis in patients with systemic lupus erythematosus. Lupus 2018; 27: 1499–1508.

51. Hu S, Wu X, Xu W, et al. Long-term efficacy and safety of sirolimus therapy in patients with lymphangioleiomyomatosis. Orphanet J Rare Dis 2019; 14: 206.

52. Wen HY, Wang J, Zhang SX, et al. Low-Dose Sirolimus Immunoregulation Therapy in Patients with Active Rheumatoid Arthritis: A 24-Week Follow-Up of the Randomized, Open-Label, Parallel-Controlled Trial. J Immunol Res 2019; 2019:7684352.

53. Lechauve C, Keith J, Khandros E, et al. The autophagy-activating kinase ULK1 mediates clearance of free α-globin in β-thalassemia. Sci Transl Med 2019; 11: eaav4881.

54. Longo F, Piolatto A, Ferrero GB, Piga A. Ineffective Erythropoiesis in beta-Thalassaemia: Key Steps and Therapeutic Options by Drugs. Int J Mol Sci 2021; 22: 7229.

55. Ricchi P, Ammirabile M, Costantini S, et al. Soluble form of transferrin receptor as a biomarker of overall morbidity in patients with non-transfusion-dependent thalassaemia: a cross-sectional study. Blood Transfus 2016; 14: 538–540.

56. Khandros E, Kwiatkowski JL. Beta Thalassemia: Monitoring and New Treatment Approaches. Hematol Oncol Clin North Am 2019; 33: 339–353.

57. Bazvand F, Shams S, Borji Esfahani M, et al. Total Antioxidant Status in Patients with Major beta-Thalassemia. Iran J Pediatr 2011; 21: 159–65.

58. Huang Y, Lei Y, Liu R, et al. Imbalance of erythropoiesis and iron metabolism in patients with thalassemia. Int J Med Sci 2019; 16: 302–310.

59. Peterson DE, O’Shaughnessy JA, Rugo HS, et al. Oral mucosal injury caused by mammalian target of rapamycin inhibitors: emerging perspectives on pathobiology and impact on clinical practice. Cancer Med 2016; 5: 1897–907.

60. Davies M, Saxena A, Kingswood JC. Management of everolimus-associated adverse events in patients with tuberous sclerosis complex: a practical guide. Orphanet J Rare Dis 2017; 12: 35.

61. Campistol JM, de Fijter JW, Flechner SM, et al. mTOR inhibitor-associated dermatologic and mucosal problems. Clinical Transplantation Clin Transplant 2010; 24: 149–56.

62. Kaplan B, Qazi Y, Wellen JR. Strategies for the management of adverse events associated with mTOR inhibitors. Transplant Rev 2014; 28: 126–133.

63. Huang J, Nguyen-McCarty M, Hexner EO, et al. Maintenance of Hematopoietic Stem Cells through Regulation of Wnt and mTOR Pathways. Nat Med 2012; 18: 1778–1785.

64. Kahan BD. Sirolimus: a new agent for clinical renal transplantation. Transplant Proc 1997; 29: 48–50.

65. Vasquez EM. Sirolimus: a new agent for prevention of renal allograft rejection. Am J Health Syst Pharm 2000; 57: 437–48.

66. Schaffer SA, Ross HJ. Everolimus: efficacy and safety in cardiac transplantation. Expert Opin Drug Saf 2010; 9: 843–54.

67. Tang CY, Shen A, Wei XF, et al. Everolimus in de novo liver transplant recipients: a systematic review. Hepatobiliary Pancreat Dis Int 2015; 14: 461–9.

68. Ji L, Xie W, Zhang Z. Efficacy and safety of sirolimus in patients with systemic lupus erythematosus: A systematic review and meta-analysis. Semin Arthritis Rheum 2020; 50: 1073–1080.

69. Li H, Ji J, Du Y, et al. Sirolimus is effective for primary relapsed/refractory autoimmune cytopenia: a multicenter study. Exp Hematol 2020; 89: 87–95.

70. Wang Q, Luo M, Xiang B, Chen S, Ji Y. The efficacy and safety of pharmacological treatments for lymphangioleiomyomatosis. Respir Res 2020; 21: 55.

71. Sasongko TH, Ismail NF, Zabidi-Hussin Z. Rapamycin and rapalogs for tuberous sclerosis complex. Cochrane Database Syst Rev 2016; 7: CD011272.

72. Graillon T, Sanson M, Campello C, et al. Everolimus and Octreotide for Patients with Recurrent Meningioma: Results from the Phase II CEVOREM Trial. Clin Cancer Res 2020; 26: 552–557.

73. Gallo M, Malandrino P, Fanciulli G, et al. Everolimus as first line therapy for pancreatic neuroendocrine tumours: current knowledge and future perspectives. J Cancer Res Clin Oncol 2017; 143: 1209–1224.

74. Manohar PM, Beesley LJ, Taylor JM, et al. Retrospective Study of Sirolimus and Cyclophosphamide in Patients with Advanced Differentiated Thyroid Cancers. J Thyroid Disord Ther 2015; 4: 188.

75. Hortobagyi GN. Everolimus plus exemestane for the treatment of advanced breast cancer: a review of subanalyses from BOLERO-2. Neoplasia 2015; 17: 279–88.

76. Merli M, Ferrario A, Maffioli M, et al. Everolimus in diffuse large B-cell lymphomas. Future Oncol 2015; 11: 373–83.

77. Motzer RJ, Escudier B, Oudard S, et al. Phase 3 trial of everolimus for metastatic renal cell carcinoma: final results and analysis of prognostic factors. Cancer 2010; 116: 4256–65.

78. Khandros E, Thom CS, D’Souza J, Weiss MJ. Integrated protein quality-control pathways regulate free α-globin in murine β-thalassemia. Blood 2012; 119: 5265–5275.

79. Liu WJ, Ye L, Huang WF, et al. p62 links the autophagy pathway and the ubiquitin-proteasome system upon ubiquitinated protein degradation. Cell Mol Biol Lett 2016; 21:29.

80. Lei Y, Liu R, Liu J, et al. Imbalance of erythropoiesis and iron metabolism in patients with thalassemia. Int J Med Sci 2019; 16: 302–310.

81. Ganz T. Erythropoietic regulators of iron metabolism. Free Radic Biol Med 2019; 133: 69–74.

