## Supplementary material for "Expression of γ-globin genes in β-thalassemia patients treated with sirolimus: results from a pilot clinical trial (Sirthalaclin)": SIRTHALACLIN-SUPP-MedRxiv-Revised-SUBF.pdf

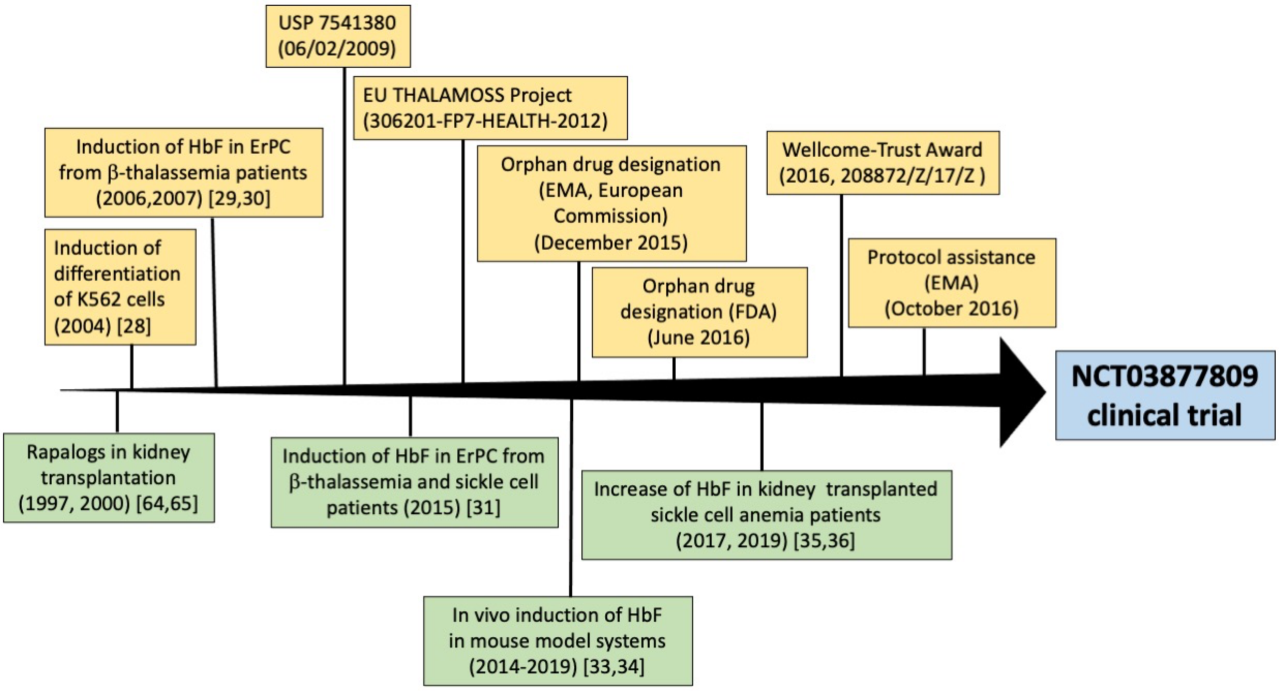

**Figure S1.** Summary of the laboratory research achievements and technological transfer activity leading to the design of the NCT03877809 trial. Yellow-boxed are activities of our group; green-boxed are key studies from the literature on sirolimus and other rapalogs.

| **TABLE S1. Inclusion criteria** | |
|---|---|
| **Age** | Patients with age over 18 years |
| **Genotype** | Patients with confirmed genotypes of homozygous/compound heterozygous β-thalassaemia major (β+/β+, β+/β0 thalassemia genotype) |
| **Clinical parameters** | Patients with documented diagnosis of transfusion dependence requiring more than 8 transfusions during the preceding 12 months |
| | Patients on regular transfusion since at least 6 years |
| | Patients with splenectomy performed at least 60 days before selection or spleen dimensions < 20 cm in the longitudinal diameter as detected by abdominal echography |
| **Management** | A medically reliable method of contraception for the entire study duration must be counseled for female participants potential childbearing |
| **Ethical/Regulatory issues** | Patients willing to follow all the study requirements and perform all the study visits and to cooperate with the investigator |
| | Patient able to understand the informed consent and to sign it before any study procedure |

| TABLE S2. Exclusion criteria | |
|---|---|
| **Analytical clinical parameters** | White blood cell count <3000 cells/ uL and/or Granulocytes <1500/ uL |
| | Platelet count <150.000/uL and >1 x106 /uL |
| | Dyslipidemia (total cholesterol > 240 mg/dl; triglycerides > 200 mg/dl) |
| | Significant proteinuria (>1g/24 hrs) |
| | Increased levels of transaminases (more than 3 times the upper limit of normal) |
| | Coexisting viral infections (positivity for human immunodeficiency virus (HIV) antibody; active hepatitis B (HBV) or hepatitis C (HCV) as demonstrated by the presence of hepatitis B surface antigen (HBsAg) and a positive HCV-RNA test, HBcAb and HBV-DNA positivity) |
| **Management/Therapy** | Patients treated with hydroxyurea at selection visit or in the last 6 months |
| | Ongoing treatment with drugs possibly affecting sirolimus actions, including treatment with Macrolidic antibiotics (chlarithromycin) |
| | Cytotoxic agents, systemic corticosteroids, immunosuppressants or anticoagulant therapy such as warfarin or heparin within 28 days before inclusion (prophylactic aspirin up to 100 mg / day is allowed) |
| | Iron chelation therapy: Deferiprone is not accepted as a chelation therapy drug, while Desferioxamina and Deferasirox are allowed |
| | Subject with any significant medical condition and/or laboratory abnormality considered by the investigator as not adequately controlled at the time of selection |
| | Treatment with live vaccines within 90 days preceding the selection |
| **Previous clinical issues** | History of severe allergic or anaphylactic reactions or hypersensitivity to excipients in the experimental drug |
| | Cardiovascular complications (heart failure as classified by the New York Heart Association (NYHA) classification 3 or higher; uncontrolled hypertension defined as systolic blood pressure (BP) ≥ 140 mm Hg or diastolic BP ≥ 90 mm Hg; significant arrhythmia requiring treatment; QTc> 450 msec on selection ECG; ejection fraction <50% by echocardiogram, MUGA or cardiac magnetic resonance; myocardial infarction within 6 months prior of selection |
| | Major surgery (including splenectomy) within 60 days before selection (patients must have fully recovered from any previous surgery) |
| | Subject with history or current malignancies (solid tumors and haematological malignancies) or presence of masses/tumor detected by ultrasound at selection |
| **Pregnancy related issues** | Pregnant or lactating women; patients who are expecting to get pregnant during the next 12 months |

| TABLE S3. Clinical data of recruited β-thalassemia patients at time of inclusion (V2) | | | | | | | | |
|---|---|---|---|---|---|---|---|---|
| | **Patient n.1** | **Patient n.2** | **Patient n.10** | **Patient n.11** | **Patient n.18** | **Patient n.21** | **Patient n.22** | **Patient n.24** |
| Average pre-transfusion Hb (g/dl) | 9.50 | 10.40 | 10.40 | 9.90 | 9.80 | 10.01 | 9.12 | 9.10 |
| Red cell consumptions (ml/kg/year) | 126.44 | 111.91 | 100.51 | 99.19 | 104.62 | 94.35 | 131.49 | 114.58 |
| White Bood Cells ($10^3$/µl) **(<3000 cells/µl)** | 12.50 | 6.71 | 17.91 | 13.94 | 12.54 | 8.17 | 8.98 | 5.80 |
| Neutrophil Granulocytes x $10^3$/µl (**<1500/ µl**) | 6.58 | 3.49 | 6.41 | 7.92 | 4.57 | 4.02 | 4.91 | 3.44 |
| Platelets x $10^3$/µl (**<150.000/ul or >1 x$10^6$/ul**) | 473 | 315 | 443 | 570 | 355 | 803 | 396 | 259 |
| Cholesterol (mg/dl) (**> 240 mg/dl**) | 189 | 147 | 139 | 164 | 125 | 168 | 121 | 94 |
| Triglycerides (mg/dl) (**> 200 mg/dl**) | 102 | 105 | 76 | 195 | 52 | 139 | 53 | 146 |
| Proteinuria (mg/ 24hrs) (**> 1g/24 hrs**) | 307 | 74 | 113 | 156 | 104 | 313 | 136 | 185 |
| Serum creatinine (mg/dl) | 0.54 | 0.79 | 0.69 | 0.61 | 0.80 | 0.75 | 0.57 | 1.02 |
| Glycemia (mg/dl) | 77 | 113 | 103 | 86 | 93 | 89 | 83 | 99 |
| Serum albumin (g/dl) | 4.34 | 4.47 | 4.35 | 4.41 | 4.19 | 4.46 | 4.18 | 4.65 |
| Transaminases (ALT-AST; UI/L)° (**< 3 times the upper limit of normal**) | 36-26 | 17-21 | 20-21 | 12-17 | 27-23 | 19-20 | 14-18 | 28-24 |
| Endocrine complications: ^ | | | | | | | | |
| Hypogonadism | yes | yes | no | yes | yes | yes | no | no |
| Hypothyroidism | no | no | no | no | no | yes | no | no |
| Hypoparathyroidism | no | no | yes | no | no | no | no | no |
| Diabetes mellitus | no | no | no | no | no | no | no | no |
| Osteoporosis | no | yes | no | yes | yes | no | yes | no |
| Iron chelation therapy: ^^ | DFX | DFX | DFO/DFX** | DFX | DFO/DFX** | DFX | DFX | DFX |
| Iron accumulation in liver and cardiac tissues (MRI-T2*) | | | | | | | | |
| Global cardiac T2* (ms)$^+$ | 37 | 37 | 42 | 41 | 36 | 35 | 34 | 37 |
| Liver iron concentration (LIC)$^{++}$ (mg(Fe)/g dry liver tissue) | 4.06 | 8.14 | 1.79 | 4.13 | 1.16 | 4.80 | 6.15 | 4.21 |

(*) Inclusion/Exclusion criteria are bolded in parenthesis
(°) ALT and AST reference range: females < 35 UI/l; males < 50 UI/L
(^) Diagnosis performed according to TIF guidelines (44)
(^^) Desferioxamine= DFO; Deferasirox = DFX; (**) sequential regimen
(+) reference range: ≥20 ms normal vaue (no significant iron load)
(++) reference range: LIC <3mg/g: no significant iron load; LIC 3-7 mg/g: mild iron load; LIC≥7 and <15 mg/g: moderate iron load; LIC ≥15 mg/g: severe iron load

All patients were negative for:

- active infection with immunodeficiency virus (HIV), hepatitis B, hepatitis C;
- treatment with hydroxyurea at selection visit or in the last 6 months;
- treatment with macrolide antibiotics (chlarithromycin), cytotoxic agents, systemic corticosteroids, immunosuppressants or anticoagulants;
- injection of live vaccines within 90 days preceding the selection;
- severe allergic or anaphylactic reactions or hypersensitivity to excipients in the experimental drug;
- cardiovascular complications, uncontrolled hypertension, significant arrhythmia requiring treatment; myocardial infarction within 6 months prior of selection;
- major surgery within 60 days before selection;
- history or current malignancies or presence of masses/tumor detected by ultrasound at selection.

| TABLE S4. Variation of blood γ-globin mRNA: relative γ-globin mRNA value (*) | | | | |
|---|---|---|---|---|
| **Patient number** | **V2** | **V6** | **V8** | **V11** |
| 1 | 12.01 | 21.02 | 18.02 | (**) |
| 2 | 7.62 | 4.36 | 15.47 | 8.61 |
| 10 | 17.71 | 38.78 | 50.83 | 48.80 |
| 11 | 1.00 | 14.05 | 13.85 | (**) |
| 18 | 3.26 | 6.72 | 12.26 | (**) |
| 21 | 1.49 | 1.94 | 0.60 | (**) |
| 22 | 7.04 | 9.15 | 30.27 | (**) |
| 24 | 16.31 | 17.94 | 13.66 | 48.59 |

(*) Values relative to patient n.11
(**) Patients 1, 11, 18, 21 and 22 concluded the trial at V8

| TABLE S5. Variation of ErPC γ-globin mRNA: relative γ-globin mRNA value (*) | | | | |
|---|---|---|---|---|
| **Patient number** | **V2** | **V6** | **V8** | **V11** |
| 1 | 4.05 | 4.86 | 3.64 | (**) |
| 2 | 1.5 | 1.05 | 5.25 | 2.25 |
| 10 | 5.5 | 6.05 | 8.25 | 15.95 |
| 11 | 1.80 | 2.70 | 10.26 | (**) |
| 18 | 1.85 | 4.44 | 3.33 | (**) |
| 21 | 4.01 | 6.40 | 2.80 | (**) |
| 22 | 2.9 | 10.73 | 4.06 | (**) |
| 24 | 1 | 7.10 | 6.50 | 1.51 |

(*) Values relative to patient n.24
(**) Patients 1, 11, 18, 21 and 22 concluded the trial at V8

| TABLE S6. Variation of total bilirubin levels (mg/dl) (**) | | | | |
|---|---|---|---|---|
| **Patient number** | **V2** | **V6** | **V8** | **V11** |
| 1 | 2.89 | 3.63 | 2.71 | (*) |
| 2 | 1.81 | 1.23 | 1.27 | 1.27 |
| 10 | 2.43 | 1.66 | 1.70 | 1.53 |
| 11 | 2.61 | 2.25 | 2.30 | (*) |
| 18 | 1.17 | 1.37 | 0.90 | (*) |
| 21 | 1.88 | 1.55 | 1.59 | (*) |
| 22 | 1.12 | 0.98 | 1.04 | (*) |
| 24 | 2.68 | 1.92 | 1.62 | 1.86 |

(*) Patients 1, 11, 18, 21 and 22 concluded the trial at V8
(**) Reference range: <1.2mg/dl

| TABLE S7. Variation of soluble transferrin receptor levels (mg/l) (**) | | | | |
|---|---|---|---|---|
| **Patient number** | **V2** | **V6** | **V8** | **V11** |
| 1 | 2.51 | 2.71 | 2.31 | (*) |
| 2 | 2.52 | 2.12 | 1.94 | 1.75 |
| 10 | 2.33 | 3.15 | 1.95 | 1.82 |
| 11 | 2.85 | 3.47 | 3.05 | (*) |
| 18 | 2.48 | 2.37 | 2.08 | (*) |
| 21 | 3.58 | 2.87 | 2.95 | (*) |
| 22 | 1.79 | 1.84 | 1.86 | (*) |
| 24 | 3.94 | 2.71 | 2.60 | 3.48 |

(*) Patients 1, 11, 18, 21 and 22 concluded the trial at V8
(**) Reference range: 0.9-2.01 mg/l

| TABLE S8. Variation of ferritin levels (ng/dl) (**) | | | | |
|---|---|---|---|---|
| **Patient number** | **V2** | **V6** | **V8** | **V11** |
| 1 | 826 | 908 | 631 | (*) |
| 2 | 1110 | 1191 | 700 | 779 |
| 10 | 842 | 546 | 499 | 488 |
| 11 | 608 | 357 | 365 | (*) |
| 18 | 552 | 545 | 414 | (*) |
| 21 | 767 | 598 | 552 | (*) |
| 22 | 542 | 504 | 477 | (*) |
| 24 | 1221 | 1213 | 1191 | 1146 |

(*) Patients 1, 11, 18, 21 and 22 concluded the trial at V8.
(**) Reference range: 11-306 ng/ml in females; 24-336 ng/ml in males

| **TABLE S9. Characteristics of stomatitis occurring during the trial** (*) | |
|---|---|
| Number of patients (%) | 5/8 (62.5%) |
| Number of episodes | 11 |
| Patients with recurrent episodes (≥2) | 3 |
| Episode duration in days, median (range) | 7.5 (1-22) |
| Time to the first stomatitis events in days, median (range) | 96 (31-190) |
| Severity (grade) (**) | Mild: 5<br>Moderate: 6 |

(*) Stomatitis or oral mucositis was defined as a disorder characterized by ulceration or inflammation of the oral mucosa. Symptoms and clinical examination: pain, superficial location, single or multiple ulcers, location in the inner lip, tongue and soft palate (dimension generally <1 cm in diameter, grayish-white colored and surrounded by an erythematous margin).
(**) From Common Terminology Criteria for Adverse Events (CTCAE) -Version 5.0 (November 27, 2017). Grade 1: asymptomatic or mild symptoms; intervention not indicated; Grade 2: moderate pain or ulcer that does not interfere with oral intake; modified diet indicated; Grade 3: severe pain; interfering with oral intake; Grade 4: life-threatening consequences; urgent intervention indicated.

| **TABLE S10. Examples of clinical studies using mTOR inhibitors** | | |
|---|---|---|
| **Clinical application** | **Drug** | **Major conclusions** |
| Organ transplantation: Kidney | Sirolimus; everolimus | Issues covered: (a) prevention of immune dysfunction and renal function preservation in de novo renal transplantation; (b) chronic dysfunction of the renal graft; (c) cardiovascular effects; (d) de novo post-transplant diabetes, and (e) de novo tumor pathology (Kahan, 1997 [64]; Vasquez, 2000 [65]). |
| Organ transplantation: Cardiac interventions | Everolimus | Everolimus may reduce late term complications, including coronary allograft vasculopathy (CAV), while maintaining the low cellular rejection rates seen with standard therapy (Schaffer and Ross, 2010 [66]). |
| Organ transplantation: Liver | Everolimus | Everolimus combined with low-dose calcineurin inhibitors (CNIs) decreases the risk acute rejection (Tang et al., 2015 [67]). |
| Lupus erythematosus (SLE) | Sirolimus | Sirolimus was found promising and well-tolerated (Ji et al., 2020 [68]). |
| Autoimmune cytopenias | Sirolimus | Sirolimus was effective for patients with primary relapsed/refractory autoimmune cytopenia with a low relapse rate and good tolerance (Li et al., 2020 [69]). |
| Lymphangioleiomyomatosis (LAM) | Sirolimus; everolimus | Sirolimus and everolimus were recommended for the treatment of LAM because they could stabilize lung function and alleviate renal AML (Wang et al., 2020 [70]). |
| Tuberous sclerosis complex | Sirolimus; Everolimus | Orally administered everolimus significantly reduced the size of sub-ependymal giant cell astrocytoma and renal angiomyolipoma (Sasongko et al., 2016 [71]). |
| Cancer: Recurrent meningioma | Everolimus | The combination of everolimus and octreotide was associated with clinical and radiological activity in aggressive meningiomas and warrants further studies (Graillon et al., 2020 [72]). |
| Cancer: Pancreatic neuroendocrine tumours (NET) | Everolimus | Everolimus should be recommended as the first line therapy for patients with symptomatic malignant unresectable insulin-secreting pNETs, to control the endocrine syndrome regardless of tumor growth (Gallo et al., 2017 [73]). |
| Cancer: Advanced Differentiated Thyroid Cancers | Sirolimus | The combination of sirolimus, with a well-known cytotoxic agent, cyclophosphamide, provides a well-tolerated and promising alternative treatment for advanced, differentiated thyroid cancers (Manohar et al., 2015 [74]). |
| Cancer: Advanced breast cancer | Everolimus | Everolimus, in combination with exemestane, is proposed for patients with advanced hormone receptor-positive/HER2-negative breast cancer (Hortobagyi, 2015 [75]). |
| Cancer: B-cell lymphomas | Everolimus | A large study on relapsed/refractory diffuse large B-cell lymphoma (DLBCL) confirmed the substantial activity and good tolerability of everolimus, with thrombocytopenia being the main toxicity. The combination of everolimus and rituximab showed encouraging results, without increasing toxicity (Merli et al., 2015 [76]). |
| Cancer: Metastatic renal cell carcinoma | Everolimus | A Phase III trial demonstrated superiority at interim analysis for everolimus over placebo in patients with metastatic renal cell carcinoma (mRCC) (Motzer et al., 2010 [77]). |

### Supplementary Methods

#### *Calculation methods for the transfusion indexes*

The period of treatment from V2 to V8 lasts approximately 180 days. However, transfusion periods only approximately correspond to the scheduled visits because transfusions were not always performed in the same day of the visit. The first transfusion episode considered as performed under therapy began in the day of V2 or in the following days. A transfusion episode includes the day of transfusion and the interval up to the following transfusion. The last transfusion episode was completed within V8 or after no more than 3 days. The transfusions performed in a period of about 180 days before V2 were considered for the estimate of the baseline blood consumption.

As an example:

| **Patient n.11** | **Duration (days)** |
|---|---|
| **Treatment with sirolimus** | 175 |
| **Period of transfusion under sirolimus** | 177 |
| **Baseline period of transfusion** | 167 |

At each transfusion, the pre-transfusion hemoglobin concentration was measured; the number of red cell concentrates transfused was recorded; the volume of pure red cells transfused was estimated from the volume and the hematocrit of the blood unit. The patients' body weight was measured regularly. Therefore, for each period of transfusion, the following parameters were calculated: (a) average pre-transfusion hemoglobin concentration (g/dL); (b) total number of red cell concentrates transfused in the period; (c) total volume of pure red cells transfused (mL); (d) average body weight (kg); (e) red cell consumption, in mL of pure red cells per kg body weight per year (this adjustment is necessary because the body weight may change from the baseline and the transfusion periods have slightly different durations); (f) index of transfusion demand is calculated dividing the red cell consumption by the average pre-transfusion hemoglobin concentration (this further adjustment is advisable because the average pre-transfusion hemoglobin concentration, too, may change from the baseline). If the endogenous production of hemoglobin increases during the treatment with sirolimus, then this parameter should decrease proportionally.
